# Changes in the profile of adults diagnosed as autistic since 2010: population based studies in the United Kingdom and Sweden

**DOI:** 10.64898/2026.05.20.26353486

**Authors:** Aws Sadik, Michael Lundberg, Golam M. Khandaker, Antonio F. Pardinas, Brian K. Lee, Paul Madley-Dowd, Cecilia Magnusson, Dheeraj Rai

## Abstract

**Objective:** To understand if sociodemographic and neuropsychiatric characteristics of people diagnosed with autism in the United Kingdom (UK) and Sweden have changed since 2010.

**Design:** Cross-context population-based cohort studies.

**Setting:** UK primary care records from 2010-2023 and Swedish population-wide register linkages from 2010-2021

**Participants:** 24,537,039 individuals age 16 or over, registered with general practices in the UK, including 141,119 with an autism diagnosis. 9,096,874 people age 16 or over in the Swedish Total Population Register, including over 100,817 with an autism diagnosis.

**Main outcome measures:** Annual age-standardised incidence and prevalence of adult autism diagnoses within different sociodemographic groups. Annual age-standardised proportion of adults with new autism diagnoses, lifetime autism diagnoses, and no autism diagnoses, with prior records of other neuropsychiatric conditions or medications.

**Results:** Incident adult autism diagnoses were consistently higher in Sweden than the UK, however incidence increased rapidly in the UK after 2020. Incident diagnoses increased fastest for 16-20-year-olds and females in both nations, as well as people in White ethnic groups in the UK and people with Swedish-born parents in Sweden. For example, in the UK in 2023 the age-standardised incidence of autism diagnoses among 16-65 years olds was 11 diagnoses per 10,000 person-years (95%CI: 10.7, 11.3) in the White ethnic group and 2.2 diagnoses per 10,000 person-years (95%CI: 1.9, 2.5) in the South Asian ethnic group. Over time there has been a consistent decline in the proportion of autistic adults with a prior diagnosis of epilepsy, psychosis and intellectual disability and an increase in the proportion with a prior diagnosis of ADHD, anxiety, depression and several other mental illnesses. For example, in the UK between 2010 and 2023 the age-standardised proportions of newly diagnosed autistic adults with prior records of epilepsy decreased from 10% (95%CI: 7.6, 13) to 4% (95%CI: 3.6, 4.5), while the proportion with records of anxiety increased from 28.7% (95%CI: 24.4, 33.6) to 58.3% (95%CI: 56.6, 60.1). Mental health conditions were generally more common in females and the reduction over time in intellectual disability was greater in females than males.

**Conclusions:** The socio-demographic and neuro-psychiatric characteristics of individuals diagnosed as autistic have changed dramatically since 2010, a phenomenon observed both in the UK and Sweden. The extent to which these changes indicate nuanced recognition of autism or broadening of diagnostic practice needs investigation.

**Summary box:** *What is already known on this topic?:* Autism diagnoses have increased internationally, with prevalence in high income countries starting to overtake estimates from population survey-based methods. It is unclear how much this reflects improved understanding of nuanced presentations or broadening of the medical concept of autism beyond coherent boundaries.

*What this study adds:* The social and medical backgrounds of people receiving autism diagnoses has changed dramatically over time, particularly for females in the UK. This suggests that: (1) diagnoses are inequitably distributed in society, (2) other mental health conditions are adding complexity to the diagnostic process, and (3) the average health needs of autistic groups are evolving with consequences for public health policy. Further research is required to understand the drivers of these changes and consequences on the provision of support for autistic people.

## Introduction

Autism is a neurodevelopmental condition characterised by social communication difficulties and restricted, repetitive patterns of behaviours and interests. The number of people being diagnosed as autistic has increased worldwide in recent decades [1][2][3], yet the distribution of autistic features across the population appears to have remained stable [4][5][6][7]. In the United Kingdom (UK) and Sweden there appeared to have been a greater increase in diagnoses of autism without language or cognitive delays compared to autism with language or cognitive delays [2][1]. There is also some evidence in Sweden that more recently diagnosed autistic children have lower autism symptom scores on average than children diagnosed in the past [8]. These findings suggest that there has been a general change in the profile of people being diagnosed with autism. This may be an appropriate shift caused by increased public awareness of autism, reduced stigma [9], greater resources available for diagnosis, and professional appreciation of more nuanced features, such as autistic presentations among women [10]. However, it has also been suggested that diagnostic practice may have become over-inclusive [11], possibly through changing views about the level of impairment associated with autistic traits [12], misattribution of symptoms of mental health conditions such as social anxiety to autism, or an autism diagnosis being seen as a way to unlock support for vulnerable individuals who would otherwise not be served by over-stretched health, education, or social care services [13]. From a health-care perspective, over-inclusive diagnostic practice could promote a pathologisation of self-identity, shift attention away from modifiable causes of distress, and potentially lead to misallocation of resources.

To understand why autism diagnoses are increasing, we can look to the sociodemographic and neuropsychiatric backgrounds of those being diagnosed. Studies in the United States of America (USA) have generally reported the prevalence of autism to be in higher families with higher socioeconomic and educational status [14][15][16]. However the inverse has been found in regions with more equitable healthcare systems, including Europe [17][18][19], Canada [20], and Japan [21]. If increasing diagnoses are focused within certain sociodemographic groups, then cultural trends or healthcare disparities may be key drivers. Neuropsychiatric diagnoses and medication histories offer added insight by illustrating diagnostic complexity and potential support needs [22]. For example, intellectual disability (ID) can be a marker of functional impairment, and “boundary conditions” such as social anxiety disorder can be difficult to differentiate from autism. Sociodemographic patterns and healthcare practice can vary between nations. Therefore, comparing trends across nations with different social patterns and healthcare services can help to clarify whether trends are generalisable or localised phenomena.

Using UK primary care records up to 2023 and Swedish population-wide register linkages up to 2021, we studied the following questions: (1) Has there been a change in the sociodemographic patterning of incident and prevalent autism diagnoses in adults? (2) Has there been a change in the frequency that other neuropsychiatric conditions are recorded in those with incident and prevalent autism diagnoses? (3) Have these trends varied by sex. Both incident and prevalent diagnoses were assessed because incident diagnoses offered contemporaneous reflections of diagnostic trends, while prevalent diagnoses integrate information from childhood and prior calendar years to outline current public health needs.

## Methods

### Data sources

In the UK we used the March 2024 extract of Clinical Practice Research Datalink (CPRD) Aurum. CPRD Aurum is a database containing routinely collected electronic health records from primary care practices that use EMIS Web [23].The March 2024 extract included over 47,413,279 research acceptable patients (as defined by CPRD) from 1,784 general practices in the UK including over 16 million patients currently contributing data with a median follow-up time of 9.55 years among current contributors [24]. 35,240,366 of these patients were eligible for linkages, such as patient postcode-level Index of Multiple Deprivation (IMD) quintiles, at the time of the release [24].

In Sweden we used linked longitudinal, total population registers including 13,345,950 people: the Total Population Register (TPR) [25], the National Patient Register (NPR)[26], the Cause of Death Register[27] and the Longitudinal Integration Database for Occupational research (LISA) [28]. The National Patient Register has coverage of psychiatric admissions and emergency department visits in Sweden from 1973, all somatic inpatient contacts from 1987, and outpatient secondary care since 2001 [26].

### Ethics and Consent Statements

For UK analyses, all procedures involving human subjects/patients were approved by the Independent Scientific Advisory Committee of CPRD (protocol number 23_002605). Individual patient consent for this study was not required as all data sent to CPRD by GP practices is pseudonymised at source, with patients able to opt-out of data sharing for research purposes. For Swedish analyses, the Swedish Ethical Review Authority (DNR 2020–05516, 2021-05958-02, and 2022-05648-02) gave ethical approval.

### Patient and Public Involvement

Two autistic advisors helped through meetings starting before the conception of the study to shape the research questions and interpret findings. This included highlighting conditions and medications of particular interest. These contributions were grounded in their lived experience, their connections to the wider autism community, and an exploration of different lenses of how autism is conceptualised in society and how improvements to the lives of autistic people will be achieved in the future. A third autistic advisor reviewed and provided comments on the manuscript. They continue to contribute to developing dissemination plans and priorities for future research.

### Study population

People entered the study on the date that all of the following criteria were met: 1st Jan 2010, their 16th birthday and, in the UK, at least 1 year of registration at a general practice. Follow-up ended at the earliest of the following: death, no longer registered in a practice (UK), migration out (Sweden) or study end date (31st December 2023 in the UK and 31st December 2021 in Sweden). In the analysis of incident diagnosis rates, participants did not contribute follow-up time after a diagnosis of autism. For the analysis of trends in neuropsychiatric history prior to a new autism diagnosis, matching was performed to ensure alignment of reference dates between index adults and comparators. Up to 4 age-, sex-, and practice (UK) or municipality (Sweden) matched non-autistic comparator adults were selected for each newly diagnosed autistic adult. For the analysis of trends in neuropsychiatric history among all autistic adults (i.e. prevalent autism diagnoses), on 31st December for each calendar year we stratified the whole study population by whether they had ever received an autism diagnosis by that date.

### Sociodemographic and neuropsychiatric variables

In the English dataset, year of birth, sex and geographical region was defined by CPRD. An ethnic group variable with six potential values (Mixed, White, South Asian, Black, Other, or Unknown) was prepared by identifying published code lists [29][30] and searching for additional SNOMED, Read and EMIS codes in the CPRD Code Browser. Patient postcode-linked 2019 English Index of Multiple Deprivation (IMD) quintiles were used as a measure of area-level deprivation [31]. In the Swedish dataset, sex, date of birth, birth country and parents’ birth countries were obtained from the TPR and parental household income quintiles and education levels were obtained from the LISA. Birth country was coded as Sweden, not Sweden or unknown. Parental countries of birth were coded as both Sweden, neither Sweden, Sweden/not Sweden or unknown. Parental household income quintiles were taken on the index person’s birth year, so that the index person’s autism status could not impact the income variable. Parental education levels were coded as compulsory, upper secondary, further education, or unknown, and were taken on the year the index person turned 15, because this variable was only available from 1990. In primary analyses, the highest level from either parent was used for household income and education levels. Individual parent values (i.e. mother or father) were used in sensitivity analyses. The term *sex* is used throughout this study because the variable labelled *gender* in CPRD data generally reflects the sex recorded at birth, based on biological characteristics, and sex at birth is recorded in the Swedish TPR.

We identified each person’s first record of the following neuropsychiatric conditions: autism, intellectual disability, attention-deficit hyperactivity disorder (ADHD), epilepsy, migraines/headaches, anxiety, depression, psychotic disorders, manic/bipolar disorders, obsessive-compulsive disorder and related conditions (OCD-related conditions), stress-related conditions (including PTSD), eating disorders, personality disorders, suicidality/self-harm, sleep disorders, and chronic fatigue. We also identified each person’s first recorded prescription (UK) or dispensation (Sweden) of seven medication classes: antipsychotics, antidepressants, antiseizure medications, opioids, gabapentinoids, melatonin, and benzodiazepines/z-drugs (sedatives). In CPRD, SNOMED, Read and EMIS codes for psychiatric conditions and medications were identified through code lists developed by AS, a clinician and checked against previous code lists with any disagreements resolved in discussion with other clinicians in the team. Previously published code lists were used for epilepsy, migraine/headache, and chronic fatigue [32]. In Swedish registers, health conditions and medications were identified by International Classification of Disease (ICD) version 8, 9, and 10 codes and Anatomical Therapeutic Chemical classification (ATC) codes, respectively, using lists developed by AS and ML.

### Statistical analyses

In each nation, the annual incidence of autism diagnoses was calculated by dividing the number of newly diagnosed adults by the person years of follow-up among those eligible to have a first autism diagnosis. The annual prevalence of autism diagnoses was calculated on 31st December of each year by dividing the number of adults with a prior autism diagnosis by the total number of adults in the study on that date. These calculations were repeated with the population stratified by age group (16-20, 21-25, 26-30, 30-40, 40-65-years-old). Annual incident and prevalent autism diagnosis rates for each sex and sociodemographic group were age-standardised to aid comparability between strata and calendar years. We performed direct standardisation using the mid-2022 age-distribution of 16-65 year old adults in the UK [33] (Table S1), due to the low numbers of diagnoses in over 65s. Parental information in Sweden was unavailable for those born prior to 1973, so these analyses were restricted to 16-35 years old to enable use of a consistent standard population from 2010 to the study end.

For incident analyses, after matching newly diagnoses autistic adults with up to 4 non-autistic comparators each, we reported counts and percentages for their distribution of index dates by time period (the autistic person’s diagnosis date: 2010-2014, 2015-2019 or 2020 to study end) and their distribution by sex, age group at diagnosis, and aforementioned sociodemographic variables. We also reported the median and interquartile range for the length of time registered in the dataset prior to the date of study inclusion.

For each condition and medication of interest, in each calendar year we calculated the age-standardised proportion of newly diagnosed autistic adults and non-autistic comparators who had ever had that condition/medication recorded by the index autism diagnosis date. We performed direct standardisation using the empirical age-distribution of adults newly diagnosed with autism from 2010-2021 in both datasets combined (Table S2). We used five-year age bands from 16 to 65 years old for standardisation as diagnoses became sparse beyond 65 years old. We repeated these analyses for all autistic adults by using 31st December as the index date for each calendar year and stratifying all adults in the whole study population by whether they had ever received an autism diagnosis before that date. We used the empirical age-distribution of autistic adults across the datasets in 2021, restricted to 16-65 year olds to ensure that there were autistic adults at each age for each calendar year (Table S3). For each condition/medication and comparator group we also calculated both the absolute and relative changes in age-standardised proportion from 2010 to 2021. Each analysis was repeated with the groups stratified by sex.

Confidence intervals for age-standardised incidence, prevalence, and proportions were calculated using Byar’s method [34] with Dobson method adjustment [35], consistent with methods recommended by the UK Department of Health & Social Care [36]. Confidence intervals for absolute differences and rate ratios were calculated using the Agresti/Caffo method [37] and the Wald/Katz-log method [38], respectively.

## Results

### Newly diagnosed autistic adults

The UK data included 155,872,147 person-years of follow-up time between 2010 and 2023, with 38,858 receiving a new autism diagnosis in this time. The Swedish dataset included 87,155,893 person-years of follow-up time between 2010 and 2021, with 50,239 receiving a new autism diagnosis during this time. Figure 1 presents the changes over time in incident autism diagnoses by sociodemographic group. Incidence rates were consistently lower in the UK than Sweden, however appeared to grow faster in the UK than Sweden over time, particularly after 2019. In both nations, incidence was highest and increased fastest for 16-20 year olds than other age groups. Incidence for males was higher than for females in 2010, but was higher for females than males by the end of the study period. Both nations saw a divergence in rates when stratified by parental background. In the UK incidence rates increased faster over time in adults from White and Mixed ethnic groups than Black, South Asian or Other ethnic groups. In Sweden rates increased faster among adults with Swedish-born parents. These divergences were greater among females than males, as presented in Figure S1. Patterns by other socioeconomic variables were more nuanced. In the UK, incidence was higher in the most deprived areas compared to the least deprived areas, throughout the study period. But when stratifying by sex, this pattern was only evident for males, and not females by the end of the study period (Figure S1). In Sweden, throughout the study period, incidence was higher among people who did not have a parent in the highest quintile of disposable income in their birth year. Sensitivity analyses demonstrated the same pattern for stratification by father’s income but not mother’s income (Figure S9). Incidence increased more over time in people who had a parent completing upper secondary or higher education than people whose parents only completed compulsory education. This pattern was replicated in sensitivity analyses that stratified by mother’s education level, but not when incidence was stratified by father’s education level.

**Figure 1:**
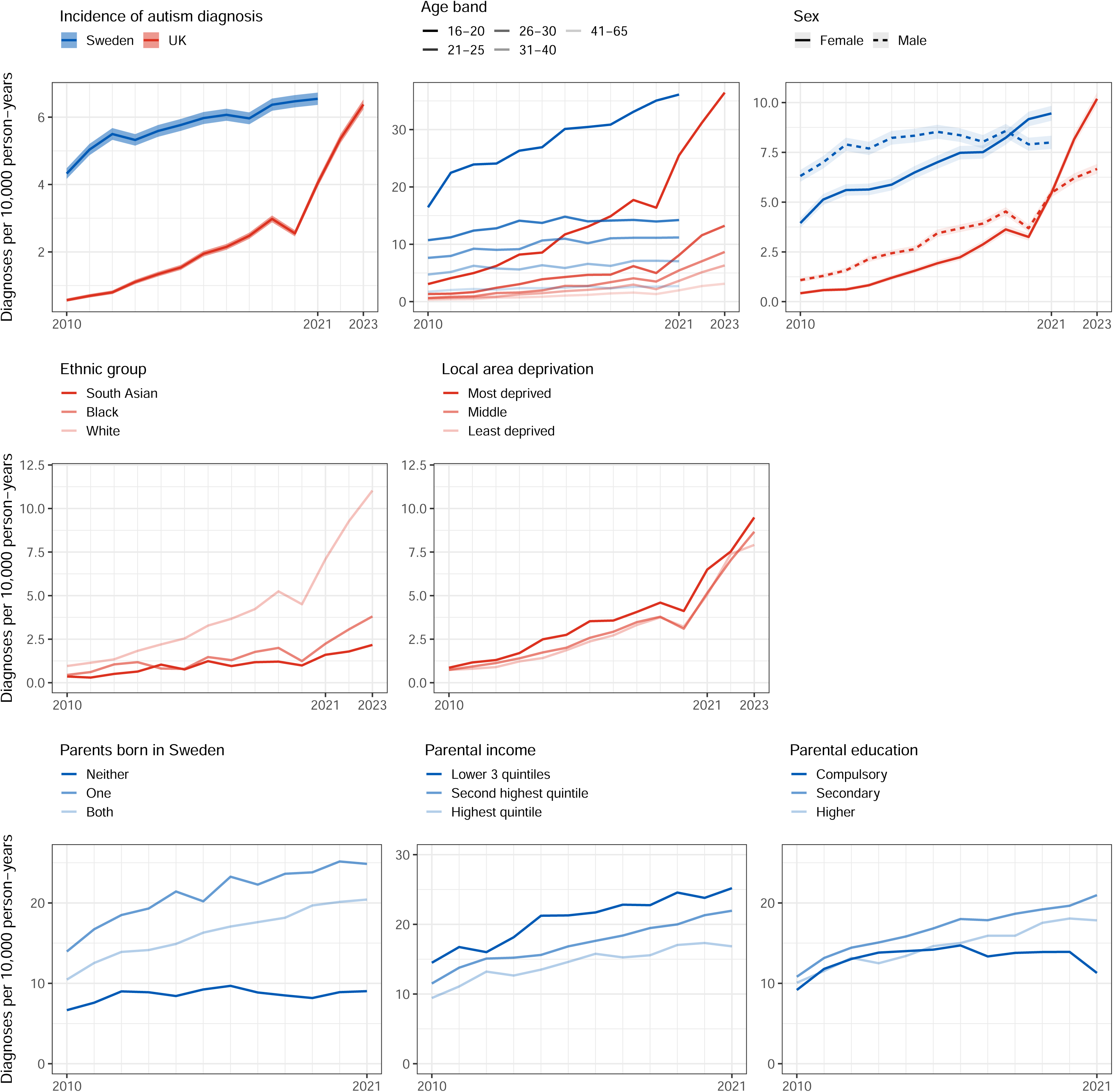
Top left: Overall annual incident autism diagnosis rates in the UK and Sweden. Top centre: Annual incidence of autism diagnosis by age group. Top right: annual age-standardised incident autism diagnosis rate by sex. Middle row: annual age-standardised incident autism diagnosis rate in the UK, by ethnic group and postcode index of multiple deprivation quintile. Bottom row: annual age-standardised incident autism diagnosis rate in Sweden, by parental birthplace, parental household income quintile at birth, and parental education at age 15. For sex, ethnic group and local area deprivation quintile, standardisation was performed using the mid-2022 age-distribution of 16-65 year old adults in the UK. For parental birthplace and education, the mid-2022 age-distribution of 16-35 year old adults in the UK was used, due to limited information for over 35s. Shaded areas = 95% confidence intervals.

For the analysis of neuropsychiatric history at the time of autism diagnosis, in the UK 38,803 of the newly diagnosed autistic adults were matched with 152,996 non-autistic adults and in Sweden 48,496 newly diagnosed autistic adults were matched with 188,421 non-autistic adults. Their sociodemographic characteristics are presented in Table 1. In the UK, autistic adults were more likely to be White and from a more deprived area than the non-autistic comparators. In Sweden, autistic adults were more likely than non-autistic adults to have a parent who completed upper secondary education and have both parents born in Sweden.

**Table 1:**
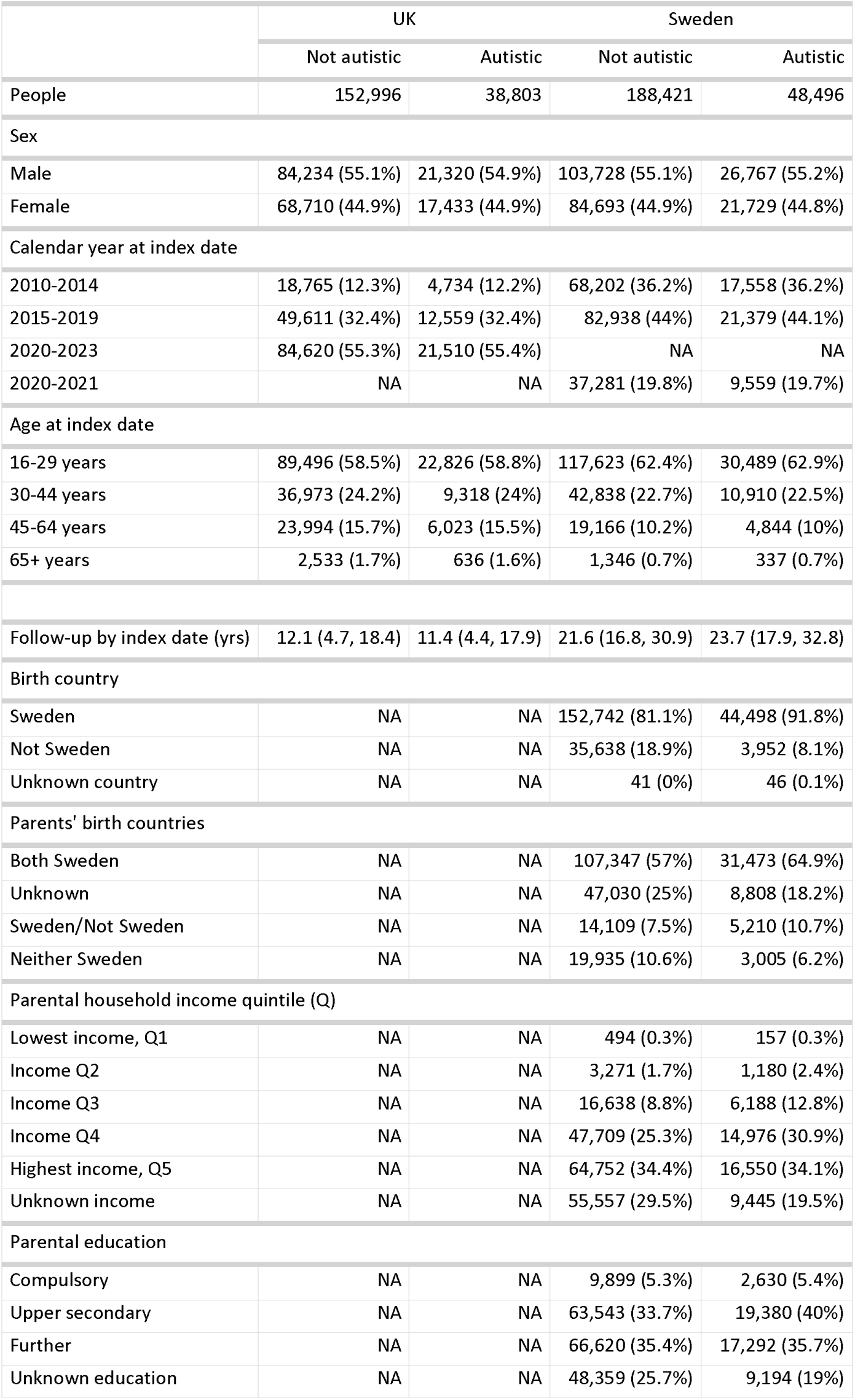
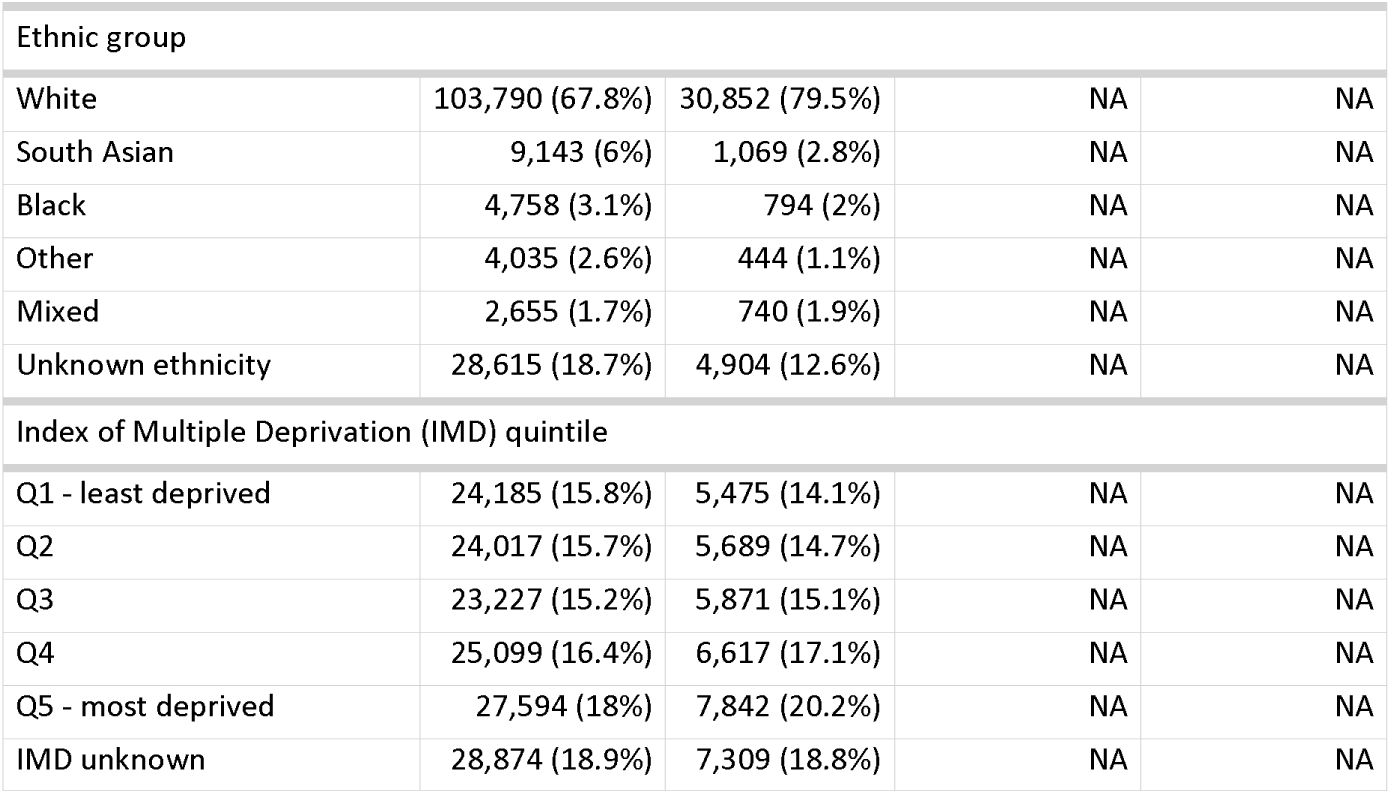
Sociodemographic characteristics of adults newly diagnosed with autism in the UK (2010-2023) and Sweden (2010-2021) and matched non-autistic comparators. Index of Multiple Deprivation quantile was calculated by patient postocde using the overall composite index for the UK in 2019.

Comparisons in neuropsychiatric history between adults diagnosed with autism in 2010 and those diagnosed in 2021 are presented in Table 2. Values for each year in the study window are presented for conditions in Figure 2 and for medications in Figure S4. Sex-stratified results for conditions are presented in Figure S2 and Figure S3. In both the UK and Sweden, more recently diagnosed autistic adults were less likely to have prior diagnoses of epilepsy, intellectual disability and psychotic disorders or prescriptions/dispensations of sedatives than those diagnosed in earlier calendar years. For example, in 2010, the age-standardised proportion of newly diagnosed autistic adults with prior records of epilepsy in the UK was 10% (95%CI: 7.6, 13) compared to 4% (95%CI: 3.6, 4.5) in 2023. These conditions did not become less common among the non-autistic comparators during the study period.

**Figure 2:**
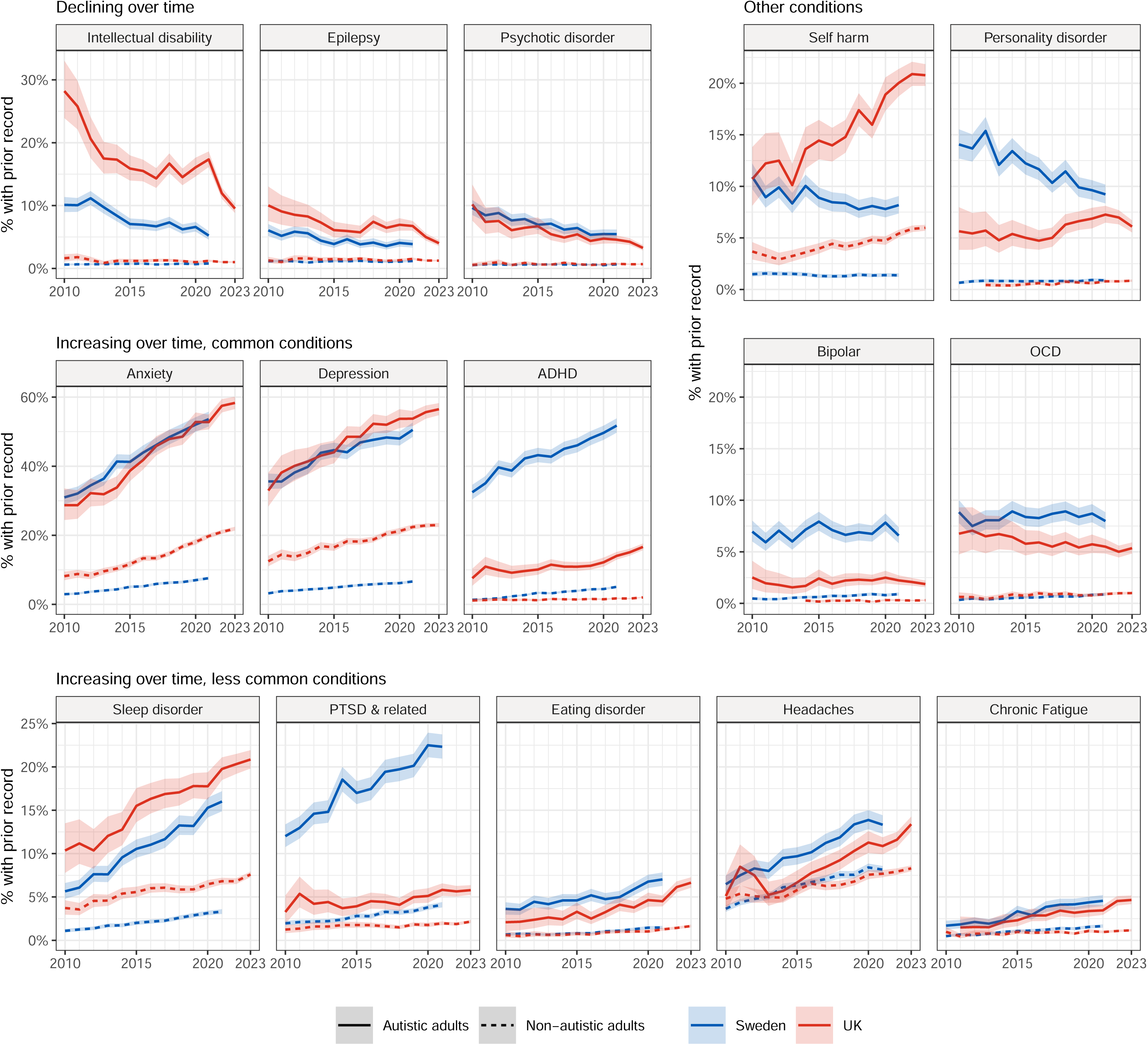
Age-standardised proportions of newly diagnosed autistic adults with prior records of neuropsychiatric conditions, by year of autism diagnosis. Direct standardisation was performed using the empirical age-distribution of adults newly diagnosed with autism across the study period in UK and Swedish datasets, using five-year age bands from 16 to 65 years old. Shaded areas = 95% confidence intervals. ADHD = attention deficit hyperactivity disorder.

**Table 2:**
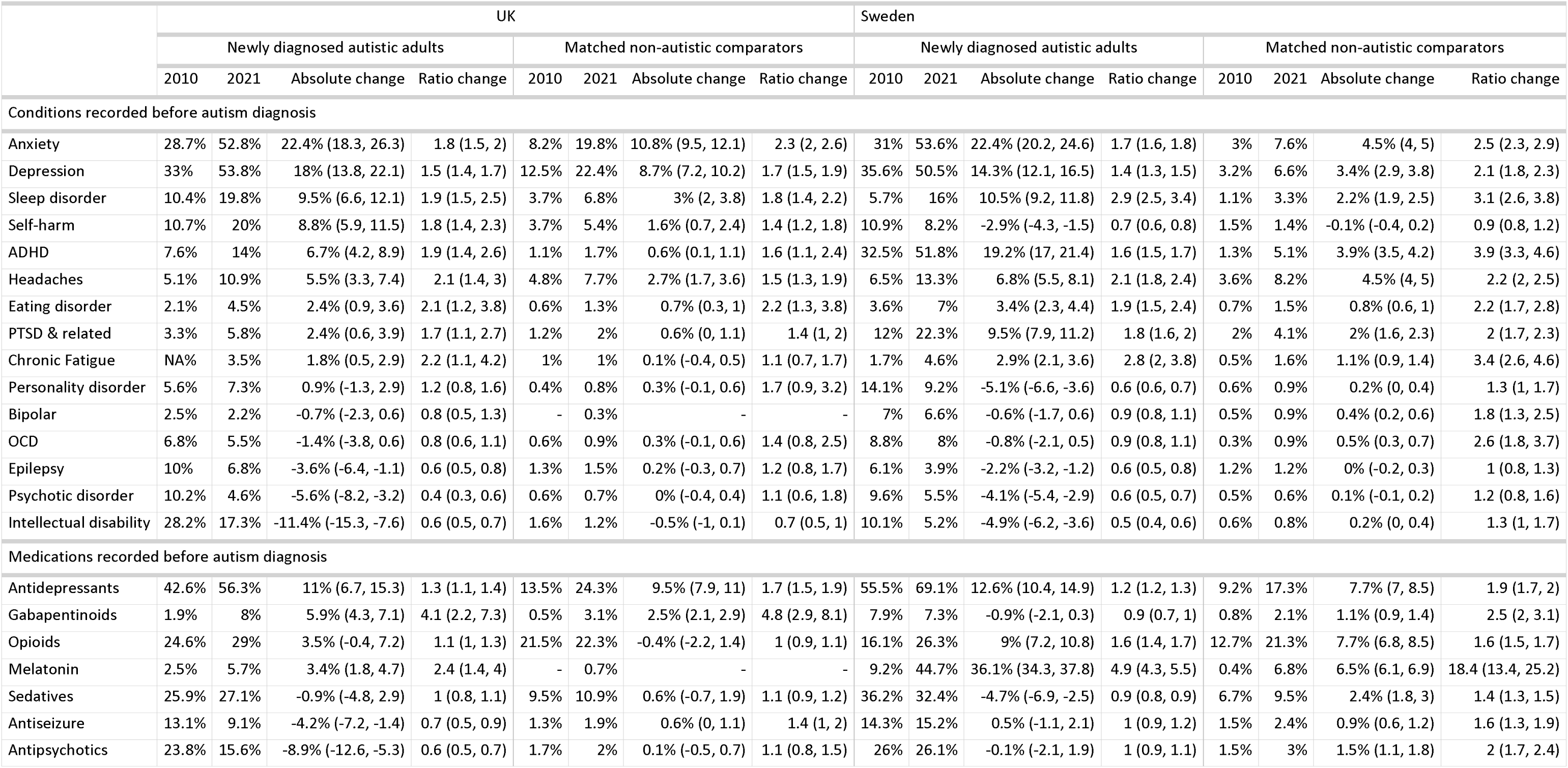
Age-standardised proportion of newly diagnosed autistic adults and non-autistic comparators in the UK and Sweden with prior records of neuropsychiatric conditions or medications in 2010 and 2021. Absolute and relative differences between 2010 and 2021 are also presented for each group, in the columns labelled Change and Ratio, respectively. Direct standardisation was performed using the empirical age-distribution of adults newly diagnosed with autism across the study period in UK and Swedish datasets, using five-year age bands from 16 to 65 years old. Confidence intervals for absolute differences and relative differences were calculated using the Agresti/Caffo method and the Wald/Katz-log method, respectively. ADHD = attention deficit hyperactivity disorder. PTSD & related = post-traumatic stress disorder and stress-related conditions.

In contrast, more recently diagnosed autistic adults were more likely to have a prior record of anxiety, depression, ADHD, sleep disorders, eating disorders, stress-related disorders, headaches, and chronic fatigue, as well as prior prescriptions/dispensation of antidepressants, melatonin, opioids, and gabapentin than those diagnosed in earlier years (Table 2, Figure 2, Figure S4). For example, the age-standardised proportion of newly diagnosed autistic adults with prior records of anxiety in the UK was 28.7% (95%CI: 24.4, 33.6) in 2010 compared to 58.3% (95%CI: 56.6, 60.1) in 2023. These conditions and medications were also more common over time in non-autistic comparators, but the absolute magnitude of increases were generally greater for autistic adults. Trends were similar in direction between females and males, however mental health conditions were generally more common for females than males and the reduction in prior records of intellectual disability was greater in magnitude for females than males (Figure S2, Figure S3).

Several trends were inconsistent between the UK and Sweden. For example, newly diagnosed autistic adults in the UK had an increasing probability over time of a prior record of self-harm, but lower probability of prescriptions for antiseizure medications, antipsychotics and sedatives. These trends were not seen, and stable, in Sweden.

### All autistic adults (prevalent autism diagnoses)

The UK data included 24,537,039 adults with follow-up time between 2010 and 2023. The Swedish data included 9,096,874 adults with follow-up time between 2010 and 2021. The prevalence patterns for autism diagnoses by sociodemographic group are presented in Figure 3, with sex-stratified results in Figure S5. In both nations, prevalence was consistently higher for males than females, increased in both groups over the study period. In the UK, the male to female ratio decrease from 4.1 in 2010, to 2.2 in 2023. In Sweden, the male to female ratio decrease from 1.9 in 2010, to 1.6 in 2021. Prevalence increased over time in all age groups and was consistently higher in younger age groups. In the UK, the prevalence of autism in 16-20 year olds was 4.5% in 2023, and in Sweden the prevalence of autism in 16-20 year olds was 3.9% in 2021. As seen for incidence, the prevalence for adults in the UK in the White ethnic group rose faster than prevalence for adults in the Black or South Asian ethnic groups, and the prevalence rose faster for adults with both parents born in Sweden than prevalence for adults with neither parent born in Sweden. Patterns for other sociodemographic variables were also broadly consistent with those seen in analyses of incidence, including for sensitivity analyses (Figure S10).

**Figure 3:**
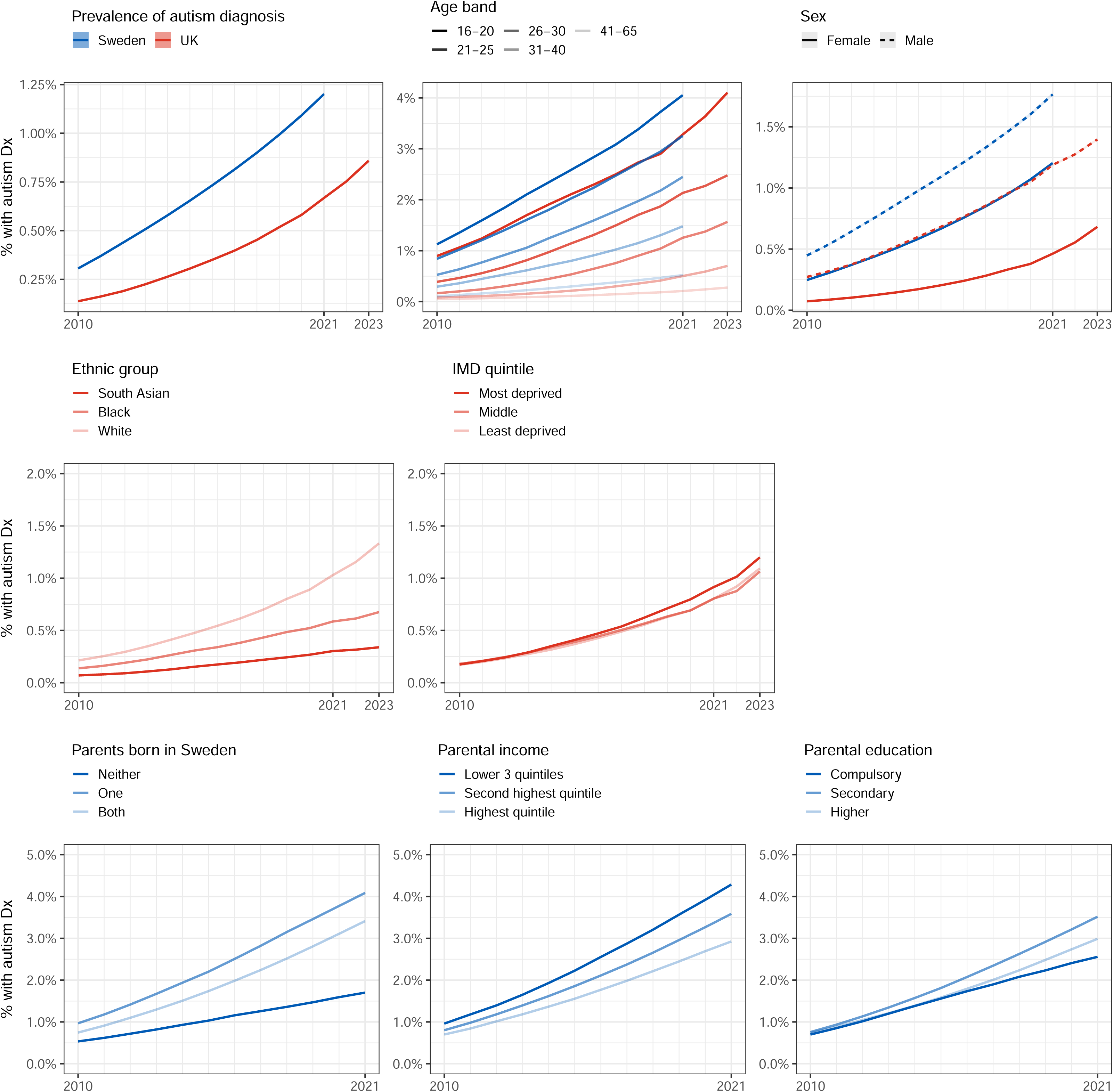
Top left: Annual autism prevalence for adults in the UK and Sweden. Top centre: Annual autism prevalence by age group. Top right: Age standardised annual autism prevalence by sex. Standardisation was performed using the mid-2022 age-distribution of 16-65 year old adults in the UK. Middle row: annual age-standardised autism diagnosis prevalence in the UK, by ethnic group and postcode index of multiple deprivation quintile Bottom row: annual age-standardised autism diagnosis prevalence in Sweden, by parental birthplace, parental household income quintile at birth, and parental education at age 15. For sex, ethnic group and local area deprivation quintile, standardisation was performed using the mid-2022 age-distribution of 16-65 year old adults in the UK. For parental birthplace and education, the mid-2022 age-distribution of 16-35 year old adults in the UK was used, due to limited information for over 35s. Shaded areas = 95% confidence intervals. Dx = diagnosis.

The time trends for past neuropsychiatric history of autistic adults are presented in Table 3, Figure 4 with sex-stratified analyses presented in Figure S6, Figure S7 and Figure S8. Changes over time and sex-differences were consistently in the same direction to those seen in the analyses of newly-diagnosed autistic adults, with decreasing ID, epilepsy and psychotic disorders and increasing anxiety, depression, sleep disorders, ADHD, stress-related disorders, eating disorders, headaches, chronic fatigue, antidepressants, melatonin, opioids, and gabapentin. For example, the age-standardised proportions of autistic adults with prior records of epilepsy in the UK decreased from 14.6% (95%CI: 13.9, 15.3) to 7.9% (95%CI: 7.7, 8.1) in 2023, while the proportion with records of anxiety increased from 18.2% (95%CI: 17.4, 19) to 44.3% (95%CI: 43.8, 44.7). All conditions, except for ADHD and psychotic disorders, were more common in autistic females than autistic males in 2010. Many conditions, including anxiety, eating disorders and chronic fatigue, increased more among autistic females than autistic males over the study period. On the other hand, ID and epilepsy decreased more among autistic females than autistic males, and, by the end of the study period, ID was more common in autistic males than autistic females.

**Figure 4:**
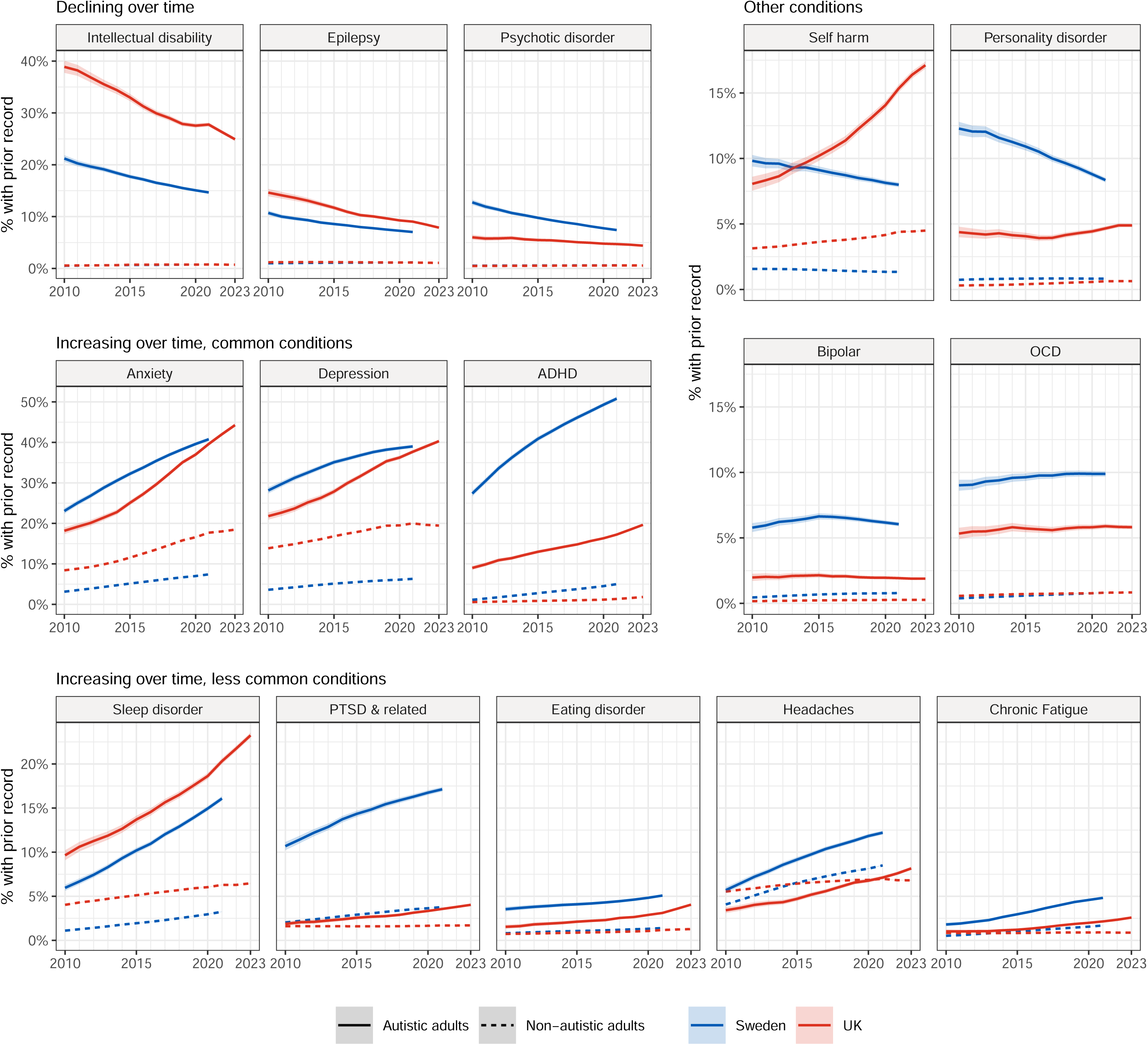
Annual age-standardised proportions of autistic adults and non-autistic adults with prior records of neuropsychiatric conditions. For standardisation we used the empirical age-distribution of autistic adults across the datasets in 2021, restricted 16-65 year olds. Shaded areas = 95% confidence intervals. ADHD = attention deficit hyperactivity disorder.

**Table 3:** Age-standardised proportion of autistic and non-autistic adults in the UK and Sweden with prior records of neuropsychiatric conditions or medications in 2010 and 2021. Absolute and relative differences between 2010 and 2021 are also presented for each group, in the columns labelled Change and Ratio, respectively. For standardisation we used the empirical age-distribution of autistic adults across the datasets in 2021, restricted 16-65 year olds. Confidence intervals for absolute differences and relative differences were calculated using the Agresti/Caffo method and the Wald/Katz-log method, respectively. ADHD = attention deficit hyperactivity disorder. PTSD & related = post-traumatic stress disorder and stress-related conditions.

|  | UK |  |  |  |  |  |  |  | Sweden |  |  |  |  |  |  |  |
| --- | --- | --- | --- | --- | --- | --- | --- | --- | --- | --- | --- | --- | --- | --- | --- | --- |
|  | Autistic adults |  |  |  | Non-autistic adults |  |  |  | Autistic adults |  |  |  | Non-autistic adults |  |  |  |
|  | 2010 | 2021 | Absolute change | Ratio change | 2010 | 2021 | Absolute change | Ratio change | 2010 | 2021 | Absolute change | Ratio change | 2010 | 2021 | Absolute change | Ratio change |
| Conditions ever recorded before year end |  |  |  |  |  |  |  |  |  |  |  |  |  |  |  |  |
| Anxiety | 18.2% | 39.6% | 22% (21.3, 22.7) | 2.3 (2.3, 2.4) | 8.4% | 17.7% | 8.6% (8.5, 8.6) | 1.7 (1.7, 1.7) | 23.1% | 40.8% | 19.5% (18.8, 20.1) | 1.9 (1.8, 1.9) | 3.1% | 7.4% | 4.1% (4.1, 4.1) | 2.3 (2.3, 2.3) |
| Depression | 21.8% | 37.7% | 16.6% (15.9, 17.4) | 1.9 (1.8, 1.9) | 13.9% | 20% | 6.6% (6.6, 6.7) | 1.3 (1.3, 1.3) | 28.1% | 39% | 13.1% (12.4, 13.7) | 1.5 (1.4, 1.5) | 3.6% | 6.3% | 3% (2.9, 3) | 1.7 (1.7, 1.7) |
| Sleep disorder | 9.6% | 20.3% | 10.5% (10, 11.1) | 2.1 (2, 2.2) | 4% | 6.3% | 3.3% (3.2, 3.3) | 1.6 (1.6, 1.6) | 6% | 16.1% | 10% (9.6, 10.4) | 2.7 (2.6, 2.9) | 1.1% | 3.2% | 2.1% (2.1, 2.1) | 2.1 (2.1, 2.1) |
| Self-harm | 8.1% | 15.3% | 7.5% (7, 8) | 2 (1.9, 2.2) | 3.1% | 4.4% | 1.1% (1.1, 1.1) | 1.3 (1.3, 1.3) | 9.8% | 8% | -0.9% (-1.4, -0.5) | 0.9 (0.9, 0.9) | 1.6% | 1.3% | 0% (-0.1, 0) | 1 (1, 1) |
| ADHD | 9% | 17.2% | 7% (6.4, 7.6) | 1.6 (1.6, 1.7) | 0.6% | 1.3% | 0.5% (0.5, 0.5) | 3.4 (3.3, 3.4) | 27.4% | 50.8% | 22.1% (21.4, 22.7) | 1.8 (1.8, 1.8) | 1.1% | 5% | 2.4% (2.4, 2.4) | 4.8 (4.7, 4.8) |
| Headaches | 3.4% | 7.1% | 3.4% (3, 3.7) | 2 (1.8, 2.2) | 5.5% | 7% | 2.3% (2.3, 2.3) | 1.4 (1.4, 1.4) | 5.7% | 12.2% | 6.7% (6.3, 7) | 2.2 (2, 2.3) | 4.1% | 8.5% | 4.9% (4.8, 4.9) | 2.2 (2.1, 2.2) |
| Eating disorder | 1.5% | 3.1% | 1.6% (1.4, 1.9) | 2.1 (1.9, 2.5) | 0.7% | 1.2% | 0.4% (0.4, 0.4) | 1.6 (1.6, 1.6) | 3.5% | 5.1% | 1.7% (1.4, 2) | 1.5 (1.4, 1.6) | 0.8% | 1.4% | 0.5% (0.5, 0.5) | 2 (2, 2.1) |
| PTSD & related | 1.9% | 3.6% | 1.6% (1.4, 1.9) | 2 (1.8, 2.3) | 1.6% | 1.7% | 0.3% (0.3, 0.3) | 1.1 (1.1, 1.1) | 10.7% | 17.1% | 8% (7.5, 8.4) | 1.8 (1.7, 1.8) | 2% | 3.8% | 2.6% (2.6, 2.7) | 2.1 (2, 2.1) |
| Chronic Fatigue | 1% | 2.1% | 1% (0.8, 1.2) | 2.1 (1.8, 2.5) | 0.8% | 0.9% | 0.3% (0.3, 0.4) | 1.2 (1.2, 1.2) | 1.8% | 4.8% | 3.1% (2.9, 3.3) | 2.8 (2.5, 3.1) | 0.5% | 1.7% | 1.3% (1.3, 1.4) | 3.5 (3.5, 3.6) |
| OCD | 5.3% | 5.9% | 0.7% (0.3, 1.1) | 1.2 (1.1, 1.2) | 0.6% | 0.8% | 0.3% (0.2, 0.3) | 1.4 (1.3, 1.4) | 9% | 9.9% | 1.4% (1, 1.8) | 1.2 (1.1, 1.2) | 0.4% | 0.8% | 0.4% (0.4, 0.4) | 2.2 (2.2, 2.3) |
| Personality disorder | 4.4% | 4.7% | 0.4% (0.1, 0.8) | 1.1 (1, 1.2) | 0.3% | 0.6% | 0.3% (0.3, 0.3) | 1.6 (1.6, 1.6) | 12.3% | 8.4% | -2.1% (-2.6, -1.6) | 0.8 (0.8, 0.9) | 0.7% | 0.8% | 0.2% (0.2, 0.3) | 1.3 (1.2, 1.3) |
| Bipolar | 2% | 1.9% | 0% (-0.2, 0.3) | 1 (0.9, 1.2) | 0.2% | 0.3% | 0.1% (0.1, 0.2) | 1.5 (1.5, 1.5) | 5.8% | 6% | 1% (0.7, 1.3) | 1.2 (1.1, 1.3) | 0.5% | 0.8% | 0.5% (0.5, 0.5) | 1.8 (1.8, 1.8) |
| Psychotic disorder | 6% | 4.7% | -0.8% (-1.2, -0.4) | 0.8 (0.8, 0.9) | 0.5% | 0.6% | 0.2% (0.2, 0.2) | 1.2 (1.2, 1.2) | 12.7% | 7.4% | -3.7% (-4.2, -3.3) | 0.7 (0.7, 0.7) | 0.5% | 0.6% | 0.1% (0.1, 0.1) | 1.1 (1.1, 1.1) |
| Epilepsy | 14.6% | 9% | -5.1% (-5.7, -4.5) | 0.6 (0.6, 0.7) | 1.2% | 1.1% | 0% (0, 0) | 1 (1, 1) | 10.7% | 7% | -3.5% (-3.9, -3) | 0.7 (0.6, 0.7) | 1% | 1.2% | 0.2% (0.1, 0.2) | 1.1 (1.1, 1.2) |
| Intellectual disability | 38.9% | 27.8% | -8.6% (-9.5, -7.8) | 0.8 (0.7, 0.8) | 0.6% | 0.8% | 0.2% (0.2, 0.2) | 1.4 (1.4, 1.4) | 21.2% | 14.7% | -6.6% (-7.2, -6) | 0.7 (0.7, 0.7) | 0.5% | 0.8% | 0.2% (0.2, 0.2) | 1.6 (1.6, 1.6) |
| Medications ever recorded before year end |  |  |  |  |  |  |  |  |  |  |  |  |  |  |  |  |
| Antidepressants | 31.8% | 46.3% | 16.1% (15.3, 16.9) | 1.6 (1.5, 1.6) | 15.4% | 22.2% | 8.7% (8.6, 8.7) | 1.4 (1.4, 1.4) | 49% | 62.2% | 15.7% (15, 16.4) | 1.3 (1.3, 1.3) | 9.6% | 17.6% | 10.5% (10.4, 10.5) | 1.8 (1.8, 1.8) |
| Melatonin | 3.5% | 10.6% | 7.3% (6.9, 7.7) | 2.7 (2.5, 2.9) | 0.1% | 0.5% | 0.3% (0.3, 0.3) | 6.5 (6.3, 6.7) | 10% | 47.1% | 35.2% (34.7, 35.7) | 4.4 (4.2, 4.5) | 0.5% | 6.7% | 5.2% (5.1, 5.2) | 11.7 (11.6, 11.9) |
| Opioids | 18.5% | 23.4% | 4.5% (3.8, 5.2) | 1.3 (1.2, 1.3) | 23.5% | 21.3% | 2.1% (2.1, 2.1) | 1.1 (1.1, 1.1) | 14.2% | 23.7% | 11.9% (11.3, 12.4) | 1.9 (1.8, 1.9) | 13.7% | 21.5% | 14.3% (14.2, 14.3) | 1.7 (1.7, 1.7) |
| Gabapentinoids | 1.7% | 5.7% | 3.5% (3.3, 3.8) | 3.4 (3, 4) | 0.8% | 2.6% | 4.2% (4.2, 4.2) | 3.4 (3.4, 3.5) | 6.3% | 7.2% | 2.1% (1.7, 2.4) | 1.4 (1.3, 1.4) | 0.9% | 2% | 2.6% (2.6, 2.6) | 2.6 (2.6, 2.6) |
| Sedatives | 27.4% | 25.3% | -1% (-1.8, -0.3) | 1 (0.9, 1) | 9.6% | 9.8% | 2.5% (2.5, 2.6) | 1.2 (1.1, 1.2) | 38.3% | 33.8% | -0.2% (-0.9, 0.5) | 1 (1, 1) | 7.1% | 9% | 4.6% (4.6, 4.6) | 1.4 (1.4, 1.4) |
| Antiseizure | 16.1% | 11.1% | -4.2% (-4.8, -3.6) | 0.7 (0.7, 0.7) | 1.3% | 1.8% | 0.7% (0.7, 0.8) | 1.4 (1.4, 1.4) | 20.1% | 18.6% | 0% (-0.6, 0.6) | 1 (1, 1) | 1.4% | 2.4% | 1.3% (1.3, 1.3) | 1.8 (1.7, 1.8) |
| Antipsychotics | 29.3% | 20.1% | -7.4% (-8.1, -6.6) | 0.7 (0.7, 0.7) | 1.4% | 1.6% | 0.2% (0.2, 0.2) | 1.1 (1.1, 1.1) | 31.9% | 32.6% | 3.1% (2.4, 3.8) | 1.1 (1.1, 1.1) | 1.5% | 2.8% | 1.6% (1.6, 1.6) | 1.8 (1.8, 1.8) |

## Discussion

This study of found dramatic changes in the sociodemographic and neuropsychiatric profiles of adults diagnosed with autism in the UK and Sweden since 2010. In both nations, incident autism diagnoses have increased most among younger adults and females, and by the 2020s new diagnoses in adult females became more frequent than for males. Prevalent autism diagnoses have also increases most among young adults, although prevalence remains higher among males than females across all adult age groups. Gaps in both incident and prevalent diagnoses have widened between ethnic groups in the UK and those with different parental backgrounds in Sweden, suggesting increasing inequality. Among both incident and prevalent autism diagnosis groups, there has also been a consistent decline over time in the proportions who have a history of intellectual disability, epilepsy and psychosis, and an increase in the proportions with a history of depression, anxiety and several other neuropsychiatric conditions. Values for most of these background characteristics varied by sex, and in the UK changes over time were generally larger in females than males.

### Comparison with other studies

A consistent increase over time in the prevalence of autism diagnoses has been reported in studies of registries and administrative records across the US and Europe. In contrast, studies based on population surveys have shown features of autism to be stable over time in the UK [7] and Sweden [6], being present in approximately 1% of the population. We found that approximately 4% of 16-20 year olds had been diagnosed with autism in both the UK and Sweden at the end of the study period, suggesting that current diagnostic practice may have broadened since 2010 and be different from the criteria used in research settings. Administrative studies in the UK and Sweden have previously reported that the relative increase in diagnoses for females has been greater than for males, resulting in declining male:female ratios [39][40]. In our current analysis, we have found that new autism diagnosis rates in adult females have overtaken those in adult males. Moreover, the male to female ratio of prevalent autism diagnoses at the end of the study period was 2.2 in the UK and 1.6 in Sweden - far lower than the 3.5:1 ratio estimated from population survey based methods[41]. The mechanism behind the widening departure from population survey-based estimates of the male to females ratio will need to be examined. For example, these findings may indicate an improved recognition of female presentations of autism, including the use of masking and camouflaging behaviours, addressing historic underdiagnosis. However, the possibility of other mental health presentations associated with camouflaging to be misidentified as autism, and in part contributing to the rapid increases in diagnosis rates among females, cannot be ruled out [42]. Indeed, the authors of the most widely used camouflaging questionnaire have recently cautioned against using the tool in making autism diagnostic decisions [43].

Associations between socioeconomic status (SES) and autism diagnosis rates have previously varied between contexts and the variables used to represent SES. US studies have previously reported lower autism prevalence in immigrant families[44], and in Black and Hispanic groups than Non-Hispanic White groups [16]. Ethnic minorities were also previously measured to have lower autism diagnoses rates in the UK [17], and in Sweden autism without ID had a lower prevalence in immigrant families [18]. In the US, patterns by ethnic group appear to have reversed over time and this has been interpreted as a sign of reducing inequalities in access to autism diagnoses within minority groups [45]. For example, in California between 1992 and 2018 diagnoses increased at faster rates for children of Black and Asian mothers [46]. However, our findings indicate that diagnoses have increased faster for those in White ethnic groups in the UK and those with parents who were both Swedish-born. This suggests that there may be cultural drivers to increased diagnoses in the UK and Sweden, and potentially increasing ethnic disparities in healthcare access. Diagnostic patterns by other markers of SES appear more complex. Regarding other markers of SES, previous US studies have generally found more autism diagnoses in higher socioeconomic status groups [14][15] [16]; while the opposite has generally been found in other high-income nations [17][20][18][19][21]. Our findings in Sweden suggest that families in which parents did not go beyond compulsory education have not had increases in autism diagnoses in line with the rest of the population. That said, autism prevalence is higher in more deprived areas in the UK and poorer families in Sweden. However, this socioeconomic patterning is no longer evident for new autism diagnoses in UK females, where instead similar diagnosis rates are seen between areas with different levels of deprivation. The reason for this is unclear, but may suggest complex sex specific dynamics that drive the process of seeking and receiving an autism diagnosis.

Previous studies in [1], the US [47], Norway [48] and Finland [19] have reported autism diagnoses to have increased more for children without co-occurring intellectual disability than those with intellectual disability. Our findings that there has been a decline in autistic adults with a past history of ID, epilepsy and psychosis is consistent with these studies, and indicates that there has been a relative increase in autism diagnoses among those with less intellectual and/or neurological impairment. It also builds on previous findings in the UK that the increase in autism diagnoses in primary care records is mostly driven by terms that are presumed to describe autism without language or cognitive delays [2]. Evidence from a Swedish twin study also showed that autism symptom scores in 7-12 year olds diagnosed with autism in 2014 were lower than for those diagnosed in 2004 [8], and a meta-analysis of 11 meta-analyses found that the differences in neurocognitive constructs between autistic and non-autistic adults appears to have declined over time [49].

A previous study in Sweden showed that more recently diagnosed autistic adults were more likely to have prior diagnoses of certain mental health conditions [22]. Our work shows that these trends: (i) extend beyond mental health conditions, to include certain medications, chronic fatigue and headaches; (ii) are generalisable across countries and data sources; (iii) are greater in magnitude than trends seen in non-autistic adults, and (iv) are also seen in adults with prevalent autism diagnoses. This latter finding emphasises that these trends are important to public health planning as well as understanding diagnostic trends. There are multiple potential explanations for the increase in mental health conditions over time. Firstly, most of these conditions have also increasingly been diagnosed among non-autistic adults in both our study and in national surveys [7] [50] - we can expect autistic adults to also be subject to these temporal trends. Secondly, changes to diagnostic manuals mean that clinicians may be more willing to recognise co-occurring conditions and diagnose autism for those with other prior mental health conditions [51]. Relatedly, it is possible that there is greater appreciation of nuanced forms of autism that may have previously been labelled as other conditions due to overlapping features, particularly in females [10]. Conversely, a greater tendency to misattribute features that are typical of other conditions to autism cannot be ruled out. Together, the trends found in this study appear to reflect the changing context that clinicians make diagnoses in [52].

The changes observed in this study have implications for planning mental health and social care support. The reasons behind the observed changes therefore need further discussion and scrutiny. Firstly, autism diagnoses have not increased as much in ethnic minority or immigrant groups, and so the mechanisms behind such disparities will need to be considered. Secondly, the high rates of previously diagnosed mental health conditions in individuals diagnosed with autism in recent years highlight the complexity in current autism diagnostic practice. Diagnostic services may need to have the skills to consider broader psychiatric formulation, including considering other diagnoses as an alternative to, or in addition to the autism diagnosis. This would contrast with the current siloed services, which are often commissioned to focus on just an autism diagnosis, and may rely on remote online methods that potentially limit assessment quality [53]. Finally, it is clear the distribution of co-occurring conditions associated with autism has changed significantly over time. Whilst these data are unable to shed light on whether these changes indicate greater appreciation of nuanced forms of autism, or misdiagnoses of other psychiatric presentations as autism, efforts will need to be made to recognise and address the needs with appropriate support.

### Strengths and limitations

Strengths of this study include the use of large prospectively-recorded population-representative datasets, reducing the risk of recall and selection bias and strengthening external validity. Also, consistent trends across two nations with different healthcare and recording practices enhances the external validity of findings. Some limitations of this study should be considered. We relied on the diagnoses and dates as recorded in the databases, and potential misclassification in these variables cannot be ruled out. Given that we sought to understand the healthcare history of those diagnosed with autism over time, rather than establish a fixed underlying estimate for the co-occurrence of neuropsychiatric conditions, such misclassification may not be a fundamental concern. Also, any misclassification of diagnosis dates is unlikely to affect the direction of trends observed over the long study period. Data for Sweden beyond 2021 was unavailable and certain findings in the UK were particularly evident after 2021, such as the decline in prior anti-seizure and antipsychotic medication prescriptions. Furthermore, the data sources in both countries have certain differences. For example, the UK data were derived from primary care records, whereas the Swedish registers primarily cover secondary care records. This may explain the contrasting trends in the results for self-harm in the two datasets: in the UK self-harm was captured if the GP had record of this, whereas in Sweden it had to be recorded during an in-patient episode. Nevertheless, the consistency of majority of the results across the two nations was reassuring.

### Conclusions

The socio-demographic and neuropsychiatric backgrounds of adults diagnosed as autistic have changed dramatically since 2010. The reasons for these changes need to be investigated in future research co-produced in partnership with the autistic community. It will be essential to do this whilst prioritising the wellbeing of autistic people and addressing health disparities in this population.

## Supporting information

Captions for supplementary tables

STROBE checklist

## Statements

## Contributor and guarantor information

The corresponding author attests that all listed authors meet authorship criteria and that no others meeting the criteria have been omitted.

## Transparency declaration

The manuscript is an honest, accurate, and transparent account of the study being reported. No important aspects of the study have been omitted. Any discrepancies from the study as planned and registered have been explained.

## Data Availability

The UK data used for this study from the Clinical Practice Research Datalink (CPRD) was obtained under licence from the UK Medicines and Healthcare products Regulatory Agency. The data is provided by patients and collected by the NHS as part of their care and support. Instructions for how to access CPRD data are available at https://www.cprd.com/access-data. The Swedish data used in this study was used under licence from Statistics Sweden. Instructions for how to access the Swedish datasets are available at https://www.scb.se/vara-tjanster/bestall-data-och-statistik/mikrodata/. The interpretation and conclusions contained in this study are those of the authors alone.

## Analytic Code Availability

Analytic code are available at https://github.com/awssadik/autism-diagnosis-changes.

## Research Material Availability

Variable code lists are available at https://github.com/awssadik/autism-diagnosis-changes.

## Acknowledgements

AS would like to thank L Ting, Ben Argo, and Sarah Douglas for their advice and feedback.

