## Supplementary material for "Changes in the profile of adults diagnosed as autistic since 2010: population based studies in the United Kingdom and Sweden": Captions for supplementary tables

### Captions for supplementary figures and tables

**Table S1:** Mid-2022 age-distribution of 16-65 year old adults in the UK. Used for direct standardisation of annual autism diagnosis incidence and prevalence estimates when stratified by sociodemographic characteristics.

**Table S2:** Age distribution of adults diagnosed with autism between 2010 and 2021 across the UK and Swedish datasets. Used for direct standardisation of annual proportions of newly diagnosed autistic adults who had prior neuropsychiatric conditions or medications.

**Table S3:** Age distribution of adults autistic adults in 2021 across the UK and Swedish datasets. Used for direct standardisation of annual proportions of all autistic adults who had prior neuropsychiatric conditions or medications.

**Table S4:** Age-standardised proportions of newly diagnosed autistic adults in the UK and Sweden with prior records of neuropsychiatric conditions, by sex and by year of autism diagnosis. Direct standardisation was performed using the empirical age-distribution of adults newly diagnosed with autism across the study period in UK and Swedish datasets, using five-year age bands from 16 to 65 years old. Shaded areas = 95% confidence intervals. ADHD = attention deficit hyperactivity disorder.

**Table S5:** Age-standardised proportions of autistic adults in the UK and Sweden with prior records of neuropsychiatric conditions, by sex and by year of autism diagnosis. For standardisation we used the empirical age-distribution of autistic adults across the datasets in 2021, restricted 16-65 year olds. Shaded areas = 95% confidence intervals. ADHD = attention deficit hyperactivity disorder.

**Table S6:** Annual incidence of autism diagnosis by age group and sex in the UK and Sweden.

**Table S7:** Annual age-standardised incident autism diagnosis rate in the UK by sex, for categories of ethnic group and postcode-level index of multiple deprivation quintile.

**Table S8:** Annual age-standardised incident autism diagnosis rate in Sweden by sex, for categories of parental birthplace, highest parental income quintile at birth, and highest parental education level at the time the index person was 15.

**Table S9:** Annual lifetime autism diagnosis prevalence by age group and sex in the UK and Sweden.

**Table S10:** Annual age-standardised lifetime autism diagnosis prevalence in the UK by sex, for categories of ethnic group and postcode-level index of multiple deprivation quintile.

**Table S11:** Annual age-standardised lifetime autism diagnosis prevalence in Sweden by sex, for categories of parental birthplace, highest parental income quintile at birth, and highest parental education level at the time the index person was 15.
