## Supplementary figures for "Changes in the profile of adults diagnosed as autistic since 2010: population based studies in the United Kingdom and Sweden"

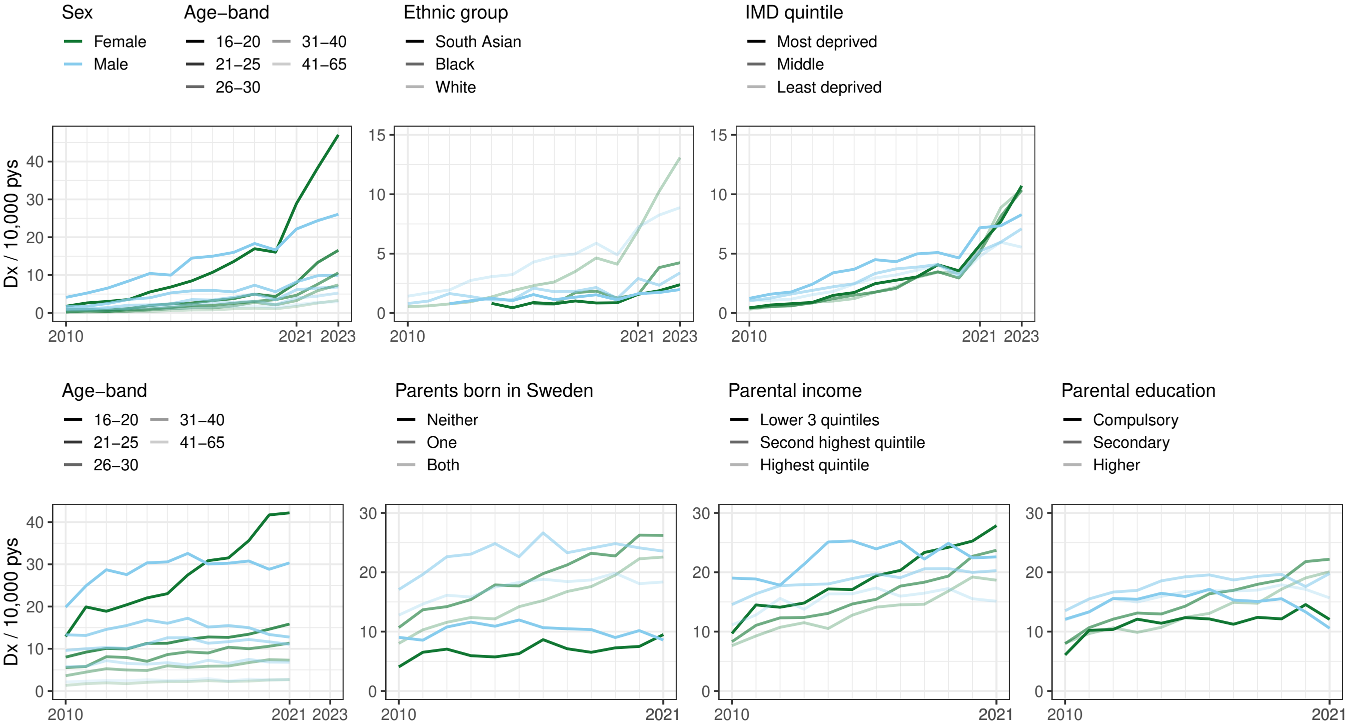


Supplementary figure 1: Top left: annual incidence of autism diagnosis by age group and sex in the UK. Top centre left: annual age−standardised incident autism diagnosis rate in the UK, by ethnic group and sex. Top centre right: annual age−standardised incident autism diagnosis rate in the UK by postcode−level index of multiple deprivation quintile. Bottom left: annual incidence of autism diagnosis by age group and sex in the UK. Bottom centre left: annual age−standardised incident autism diagnosis rate in Sweden, by parental birthplace and sex. Bottom centre right: annual age−standardised incident autism diagnosis rate in Sweden, by parental household income quintile and sex. Bottom right: annual age−standardised incident autism diagnosis rate in Sweden, by parental education and sex. For sex, ethnic group and local area deprivation quintile, standardisation was performed using the mid−2022 age−distribution of 16−65 year old adults in the UK. For parental birthplace and education, the mid−2022 age−distribution of 16−35 year old adults in the UK was used, due to limited information for over 35s.

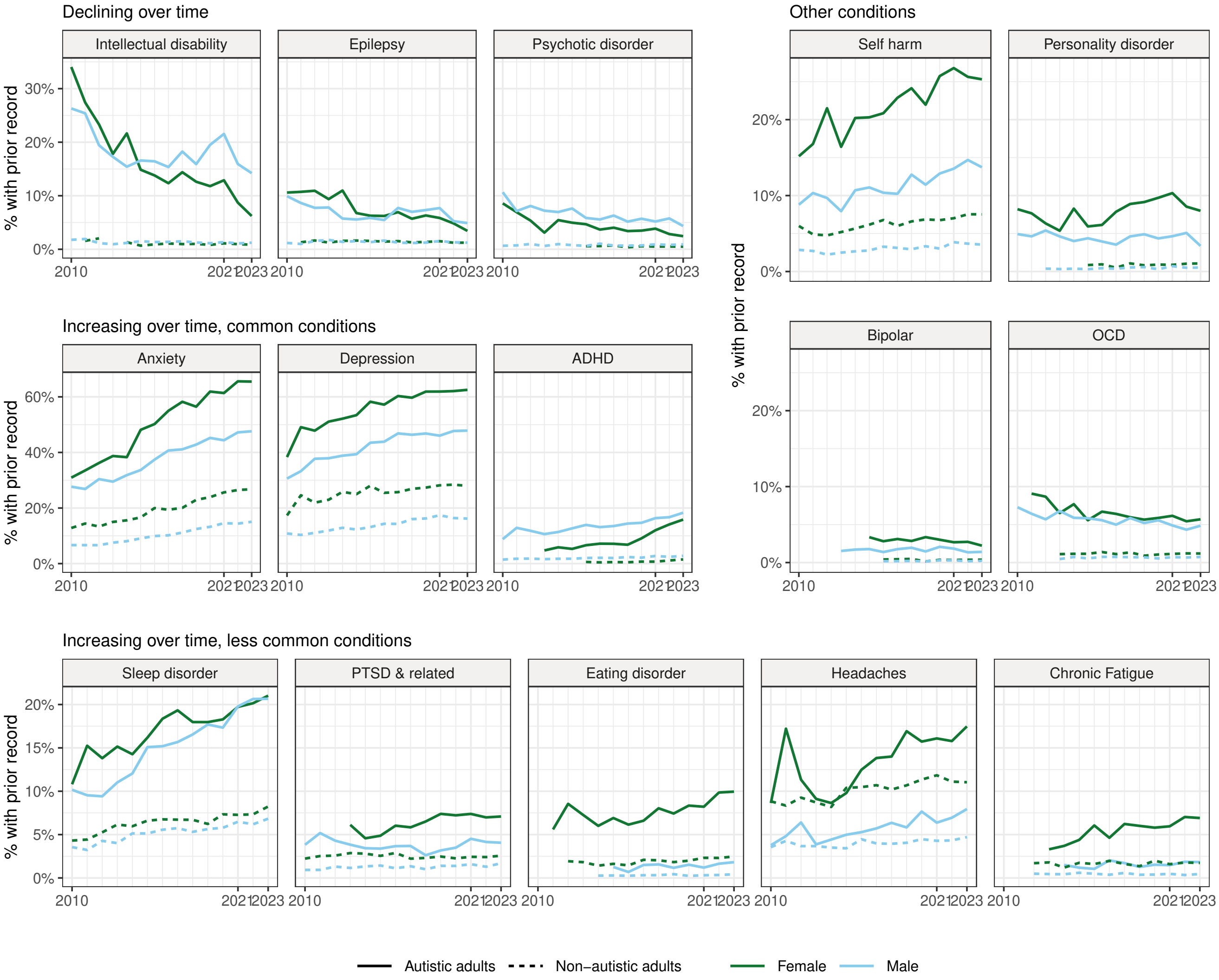


Supplementary figure 2: Age−standardised proportions of newly diagnosed autistic adults in the UK with prior records of neuropsychiatric conditions, by sex and by year of autism diagnosis. Direct standardisation was performed using the empirical age−distribution of adults newly diagnosed with autism across the study period in UK and Swedish datasets, using five−year age bands from 16 to 65 years old. Shaded areas = 95% confidence intervals. ADHD = attention deficit hyperactivity disorder.

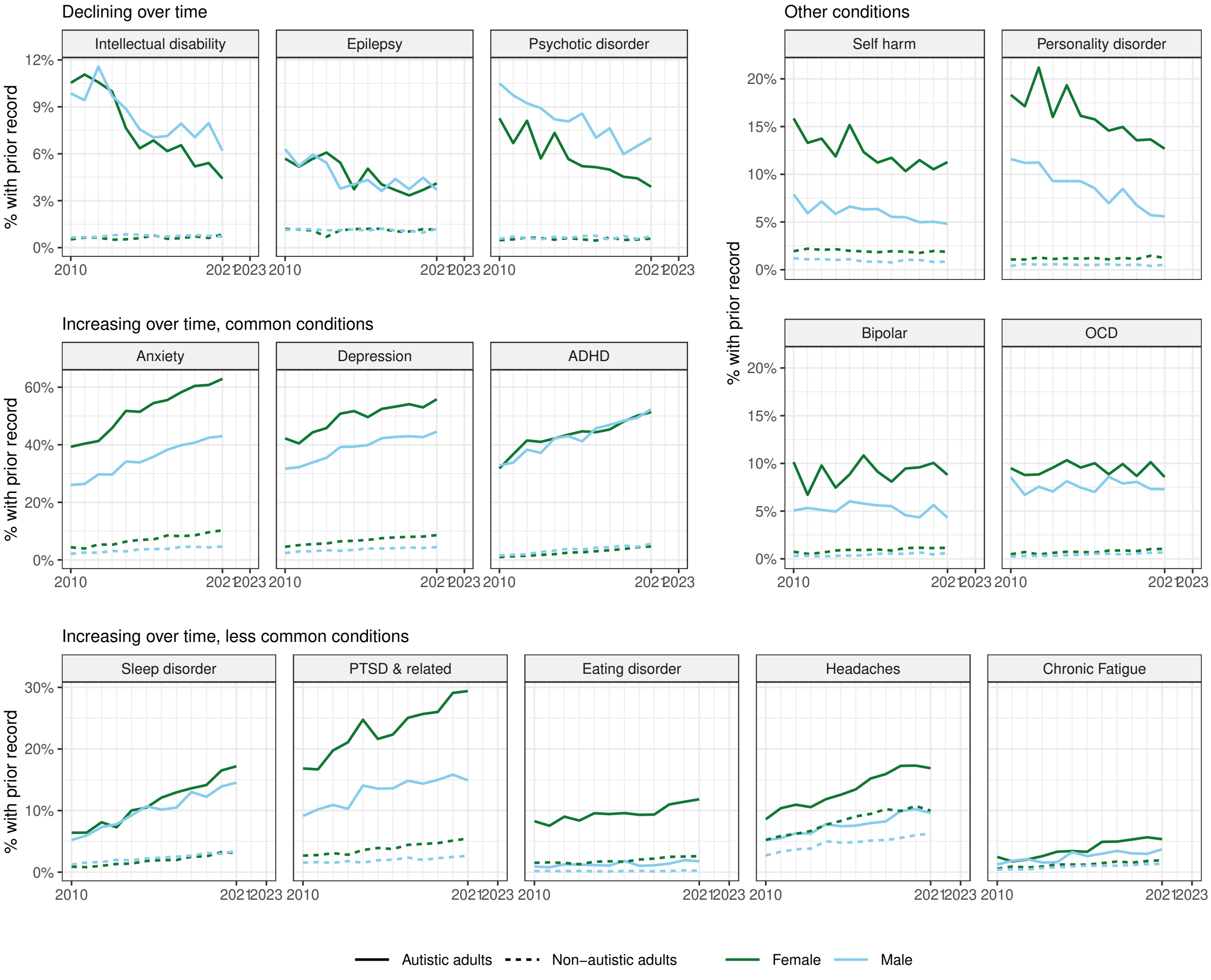


Supplementary figure 3: Age−standardised proportions of newly diagnosed autistic adults in Sweden with prior records of neuropsychiatric conditions, by sex and by year of autism diagnosis. Direct standardisation was performed using the empirical age−distribution of adults newly diagnosed with autism across the study period in UK and Swedish datasets, using five−year age bands from 16 to 65 years old. Shaded areas = 95% confidence intervals. ADHD = attention deficit hyperactivity disorder.

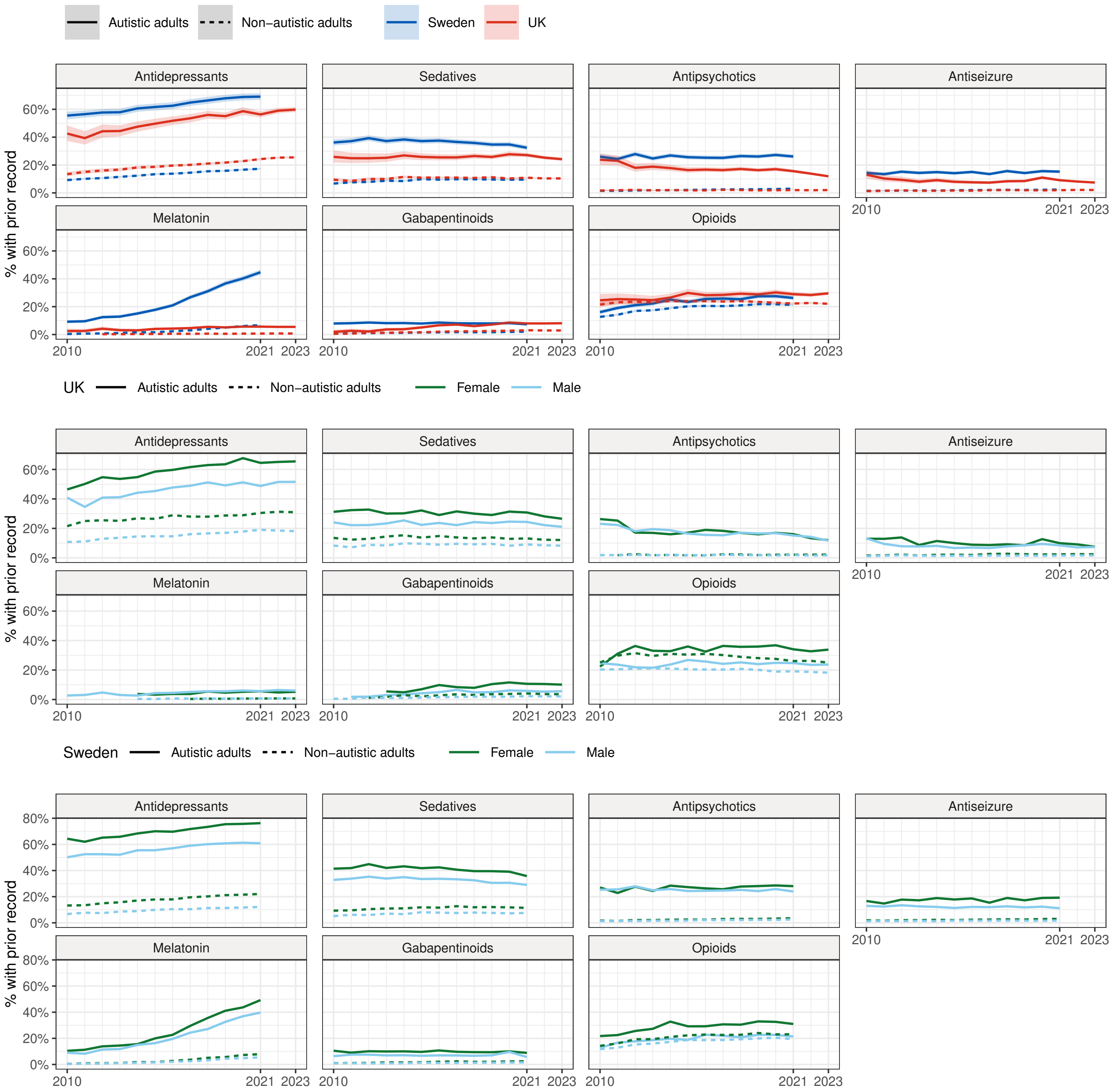


Supplementary figure 4: Top row: Age−standardised proportions of newly diagnosed autistic adults with prior records of neuropsychiatric medications, by year of autism diagnosis. Middle row: Age−standardised proportions of newly diagnosed autistic adults in the UK with prior records of neuropsychiatric medications, by sex and by year of autism diagnosis. Bottom row: Age−standardised proportions of newly diagnosed autistic adults in Sweden with prior records of neuropsychiatric medications, by sex and by year of autism diagnosis. Direct standardisation was performed using the empirical age−distribution of adults newly diagnosed with autism across the study period in UK and Swedish datasets, using five−year age bands from 16 to 65 years old. Shaded areas = 95% confidence intervals.

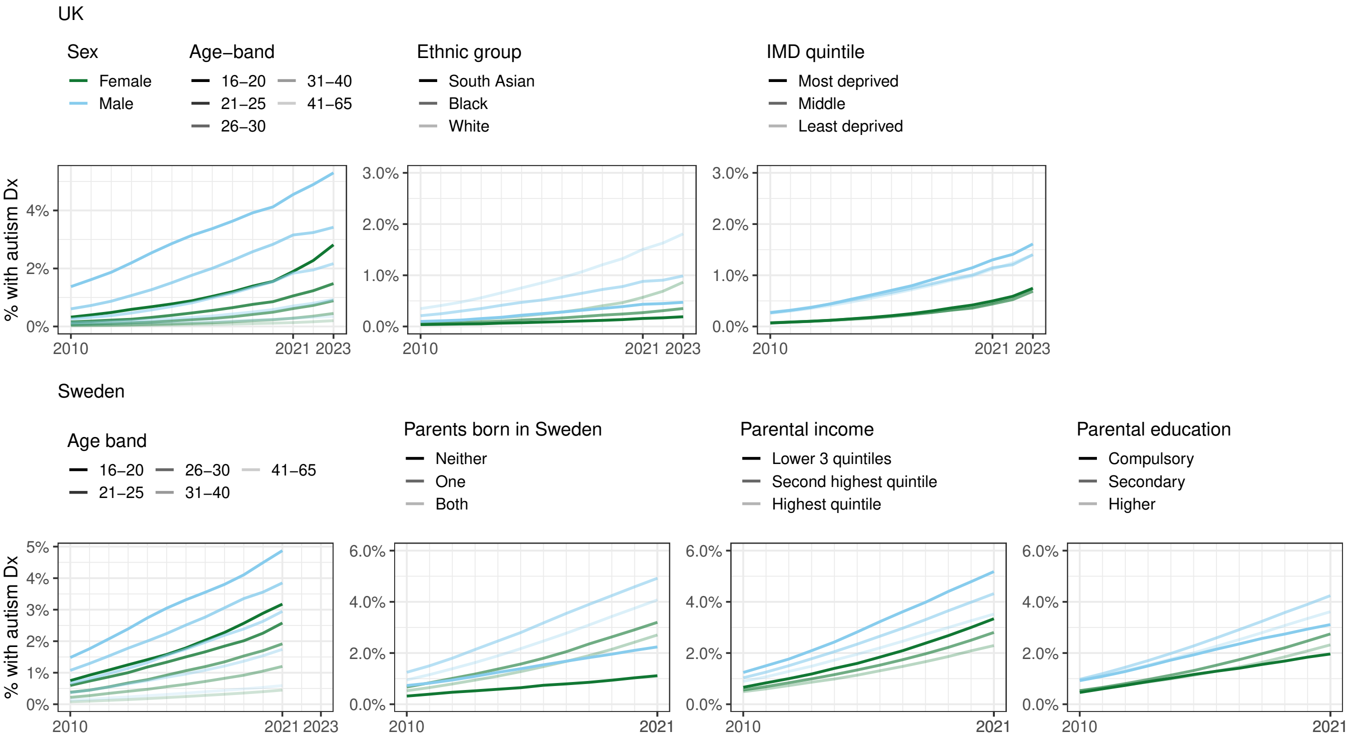


Supplementary figure 5: Top left: annual prevalence of autism diagnosis by age group and sex in the UK. Top centre left: annual age−standardised prevalent autism diagnosis rate in the UK, by ethnic group and sex. Top centre right: annual age−standardised prevalent autism diagnosis rate in the UK by postcode−level index of multiple deprivation quintile. Bottom left: annual prevalence of autism diagnosis by age group and sex in the UK. Bottom centre left: annual age−standardised prevalent autism diagnosis rate in Sweden, by parental birthplace and sex. Bottom centre right: age−standardised prevalent autism diagnosis rate in Sweden, by parental income quintile and sex. Bottom right: annual age−standardised prevalent autism diagnosis rate in Sweden, by parental education and sex. For sex, ethnic group and local area deprivation quintile, standardisation was performed using the mid−2022 age−distribution of 16−65 year old adults in the UK. For parental birthplace and education, the mid−2022 age−distribution of 16−35 year old adults in the UK was used, due to limited information for over 35s.

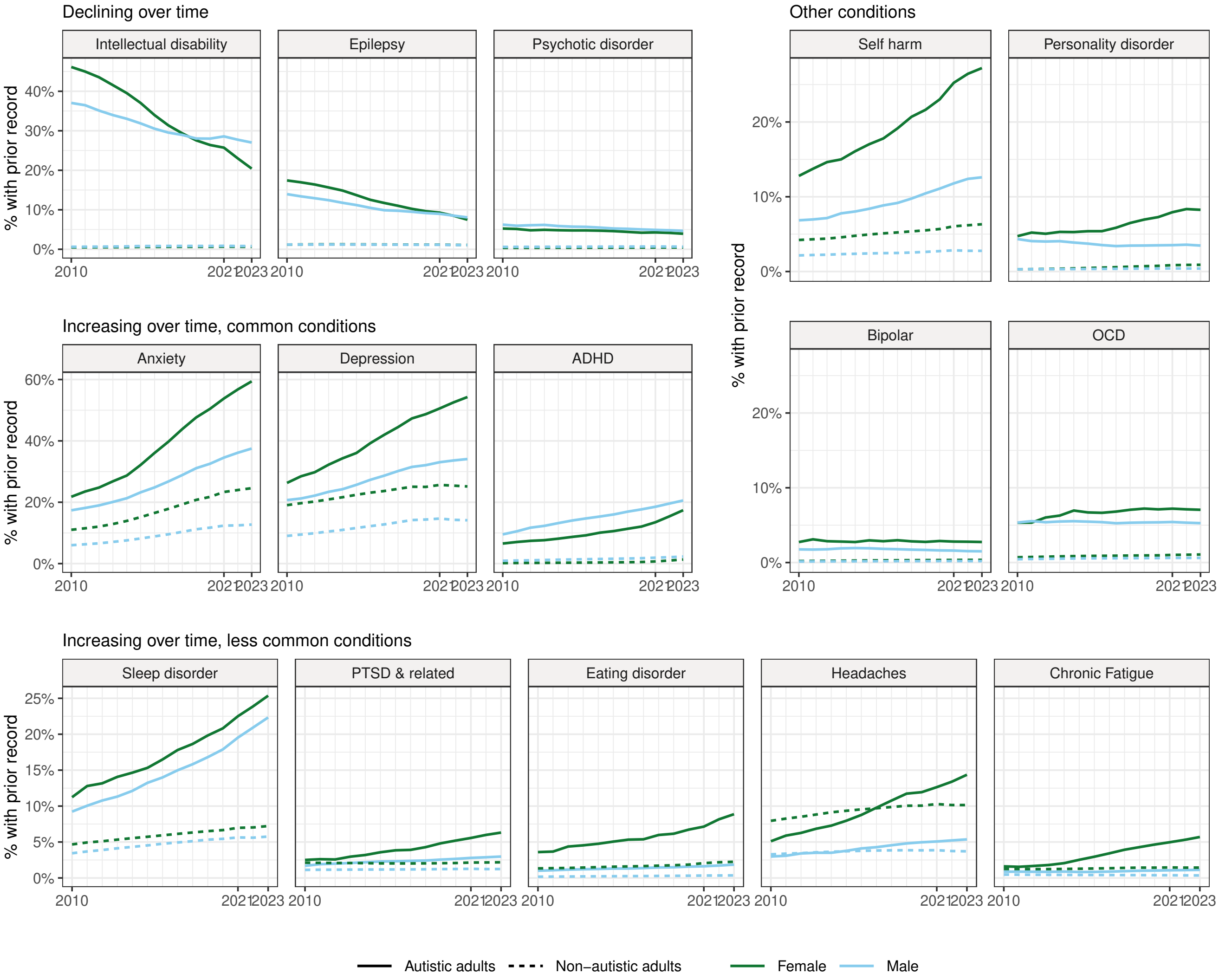


Supplementary figure 6: Age−standardised proportions of autistic adults in the UK with prior records of neuropsychiatric conditions, by sex and by year of autism diagnosis. For standardisation we used the empirical age−distribution of autistic adults across the datasets in 2021, restricted 16−65 year olds. Shaded areas = 95% confidence intervals. ADHD = attention deficit hyperactivity disorder.

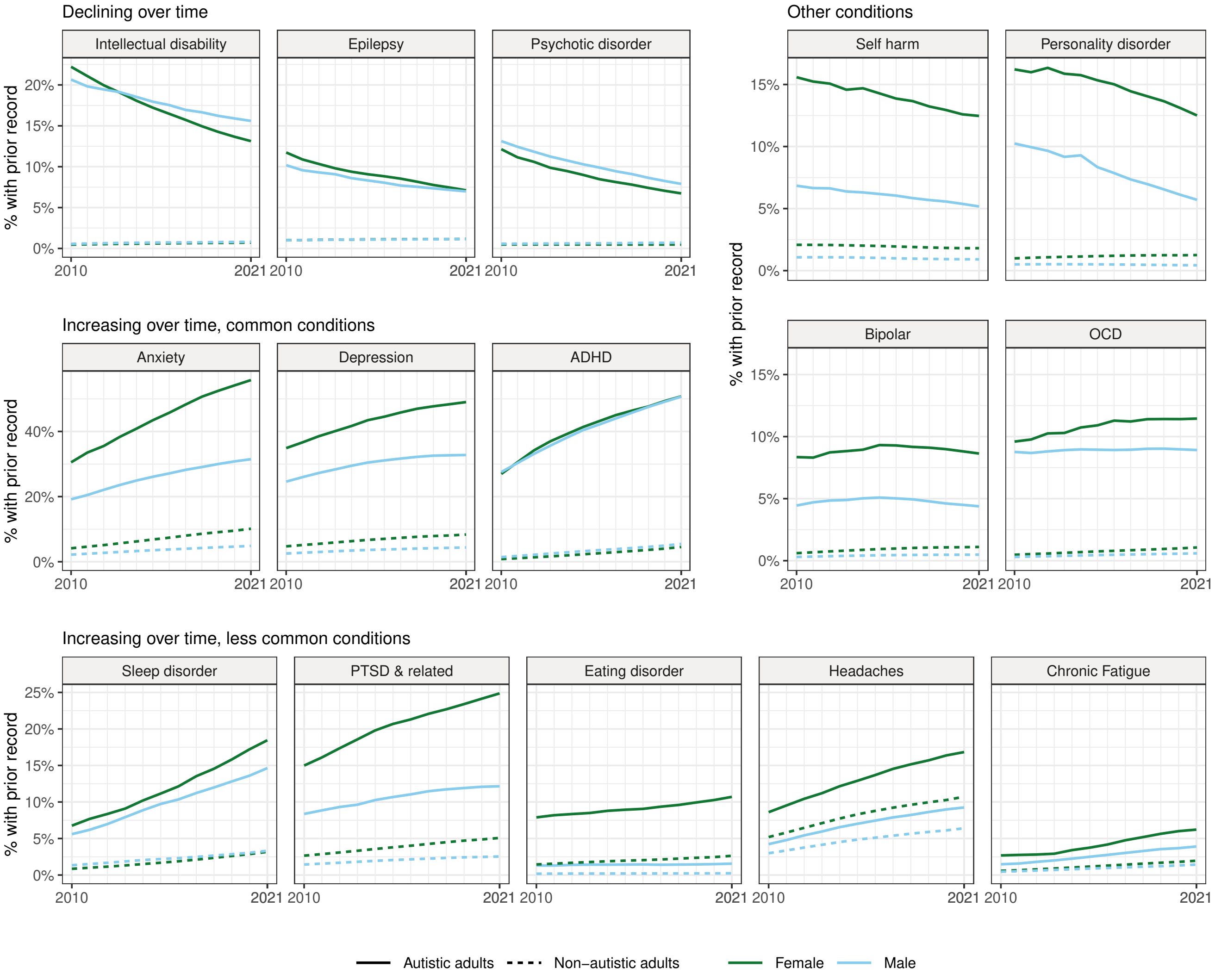


Supplementary figure 7: Age−standardised proportions of autistic adults in Sweden with prior records of neuropsychiatric conditions, by sex and by year of autism diagnosis. For standardisation we used the empirical age−distribution of autistic adults across the datasets in 2021, restricted 16−65 year olds. Shaded areas = 95% confidence intervals. ADHD = attention deficit hyperactivity disorder.

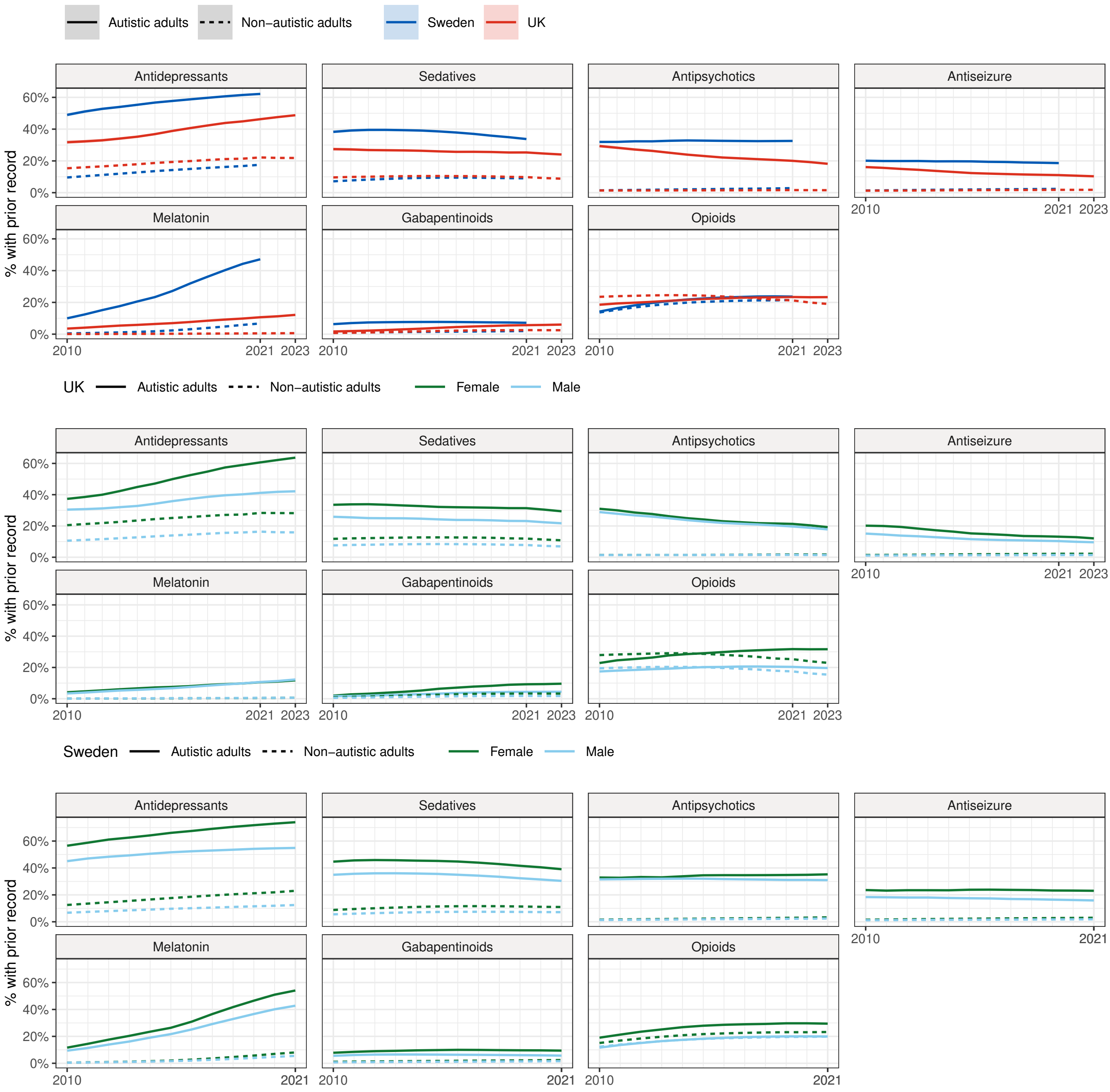


Supplementary figure 8: Top row: Age−standardised proportions of newly diagnosed autistic adults with prior records of neuropsychiatric medications, by year of autism diagnosis. Middle row: Age−standardised proportions of newly diagnosed autistic adults in the UK with prior records of neuropsychiatric medications, by sex and by year of autism diagnosis. Bottom row: Age−standardised proportions of newly diagnosed autistic adults in Sweden with prior records of neuropsychiatric medications, by sex and by year of autism diagnosis. Direct standardisation was performed using the empirical age−distribution of adults newly diagnosed with autism across the study period in UK and Swedish datasets, using five−year age bands from 16 to 65 years old. Shaded areas = 95% confidence intervals.

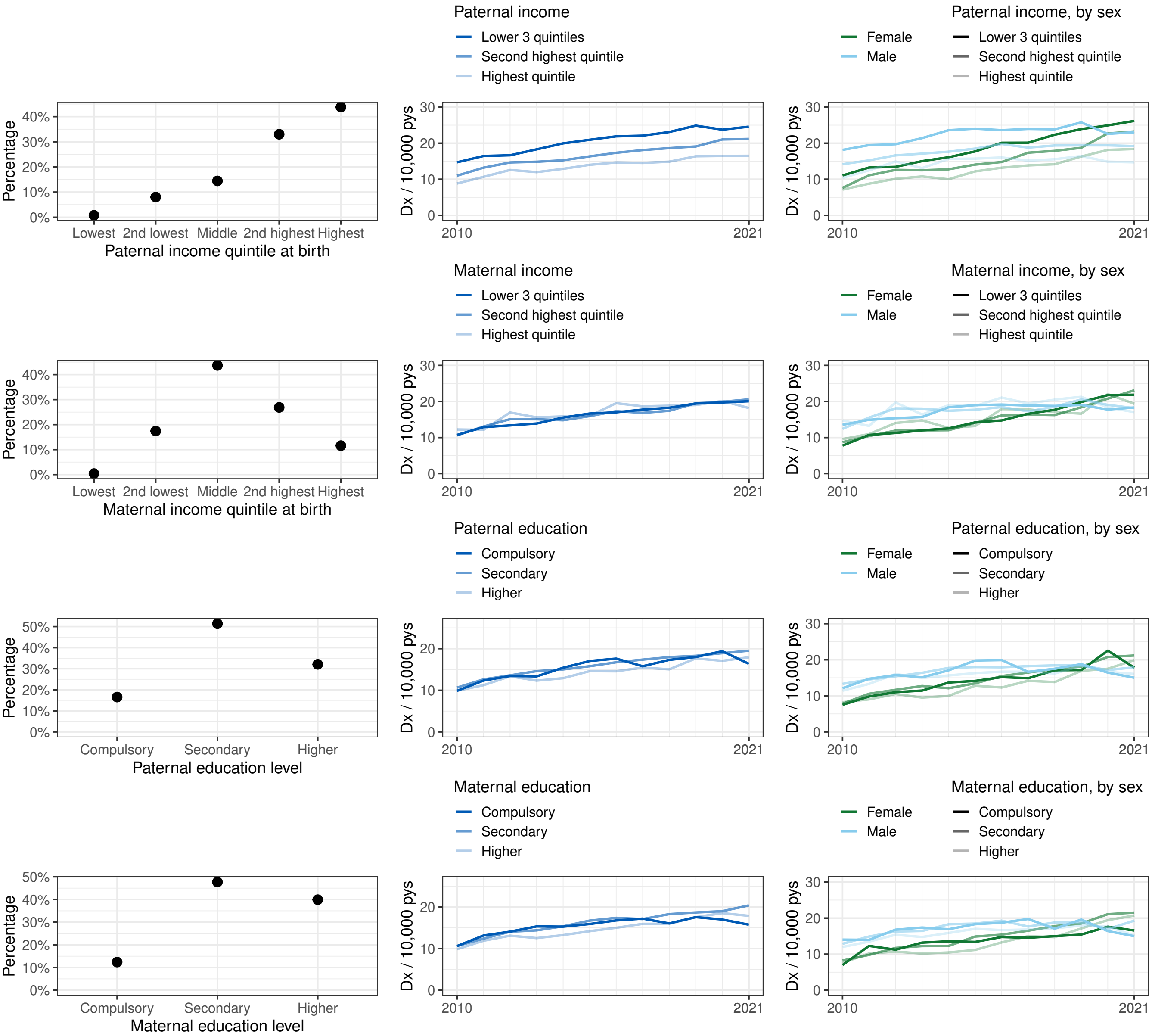


Supplementary figure 9: Top left: distribution of paternal household income quintile in adults with follow−up in 2021. Top centre: annual age−standardised incident autism diagnosis rate in Sweden, by paternal income quintle in index person's birth year. Top right: annual age−standardised incident autism diagnosis rate in Sweden, by maternal income quintle. Second row left: distribution of maternal household income quintile in adults with follow−up in 2021. Second row centre: sex−stratified annual age−standardised incident autism diagnosis rate in Sweden, by paternal income quintle in index person's birth year. Second row right: sex−stratified annual age−standardised incident autism diagnosis rate in Sweden, by maternal income quintle. Third row left: distribution of paternal education level in adults with follow−up in 2021. Third row centre: annual age−standardised incident autism diagnosis rate in Sweden, by paternal education level when index person turned 15 years old. Third row right: annual age−standardised incident autism diagnosis rate in Sweden, by maternal education level. Bottom left: distribution of maternal education level in adults with follow−up in 2021. Bottom centre: sex−stratified annual age−standardised incident autism diagnosis rate in Sweden, by paternal education level. Bottom right: sex−stratified annual age−standardised incident autism diagnosis rate in Sweden, by maternal education level. Standardisation was performed using the mid−2022 UK age−distribution of 16−35 year old adults, as parental linkage was unavailable for people born before 1973. Shaded areas = 95% confidence intervals.

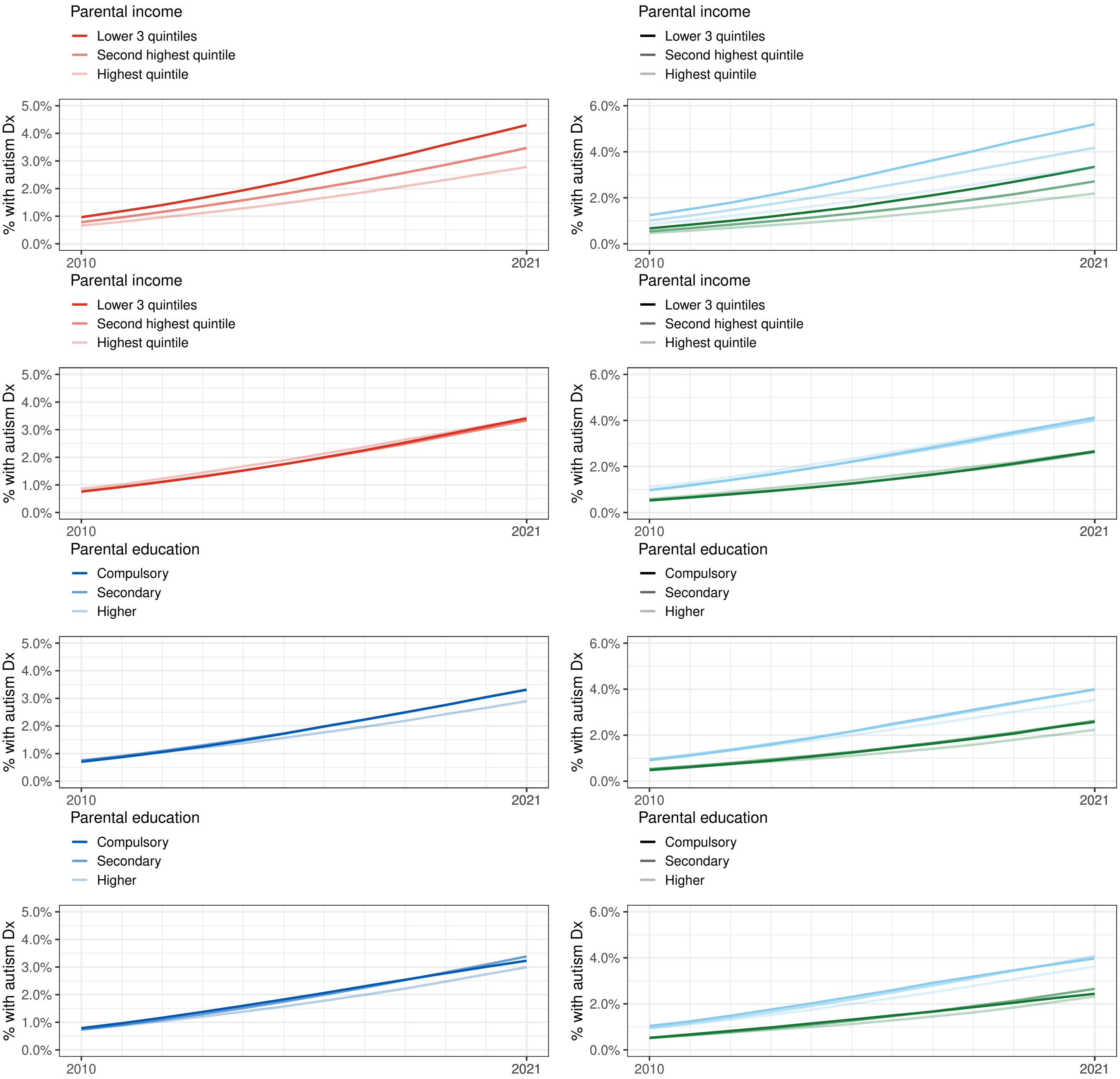


Supplementary figure 10: Top left: annual age−standardised autism diagnosis prevalence in Sweden, by paternal income quintle in index person's birth year. Top right: annual age−standardised autism diagnosis prevalence in Sweden, by maternal income quintle. Second row left: sex−stratified annual age−standardised autism diagnosis prevalence in Sweden, by paternal income quintle in index person's birth year. Second row right: sex−stratified annual age−standardised autism diagnosis prevalence in Sweden, by maternal income quintle. Third row left: annual age−standardised autism diagnosis prevalence in Sweden, by paternal education level when index person turned 15 years old. Third row right: annual age−standardised incident autism diagnosis rate in Sweden, by maternal education level. Bottom left: sex−stratified annual age−standardised autism diagnosis prevalence , by paternal education level. Bottom right: sex−stratified annual age−standardised autism diagnosis prevalence, by maternal education level. Standardisation was performed using the mid−2022 UK age−distribution of 16−35 year old adults, as parental linkage was unavailable for people born before 1973. Shaded areas = 95% confidence intervals.
